# Subgroups of young type 2 diabetes in India reveal insulin deficiency as a major driver

**DOI:** 10.1101/2021.05.07.21256703

**Authors:** Rashmi B Prasad, Olof Asplund, Sharvari R Shukla, Rucha Wagh, Pooja Kunte, Dattatrey Bhat, Malay Parikh, Meet Shah, Sanat Phatak, Annemari Käräjämäki, Anupam Datta, Sanjeeb Kakati, Tiinamaija Tuomi, Banshi Saboo, Emma Ahlqvist, Leif Groop, Chittaranjan S Yajnik

## Abstract

**Aim/Hypothesis:** Five subgroups were described in European diabetes patients using a data driven machine learning approach on commonly measured variables. We aimed to test the applicability of this phenotyping in Indian young-onset type 2 diabetes patients.

**Methods:** We applied the European derived centroids to the Indian type 2 diabetes patients diagnosed before 45 years of age from the WellGen (n = 1612) cohort. We also applied *de novo* k-means clustering to the WellGen cohort to validate the subgroups. We then compared clinical and metabolic-endocrine characteristics and the complication rates between the subgroups. We also compared characteristics of the WellGen subgroups with those of two young European cohorts ANDIS (n= 962) and DIREVA (n=420). Subgroups were also assessed in two other Indian cohorts, Ahmedabad (n = 187) and PHENOEINDY-2 (n = 205).

**Results:** Both Indian and European young type 2 diabetes patients were predominantly classified into severely insulin-deficient (SIDD) and mild obesity-related (MOD) subgroups, while the severely insulin-resistant (SIRD) and mild age-related (MARD) subgroups were rare. In WellGen, SIDD (53%) was more common than MOD (38%), contrary to figures in Europeans (Swedish: 26% vs 68%, Finnish: 24% vs 71% respectively). A higher proportion of SIDD compared to MOD was also seen in Ahmedabad (57% vs 33%) and in PHENOEINDY-2 (67% vs 23%). Both in Indians and Europeans, the SIDD subgroup was characterized by insulin deficiency and hyperglycemia, MOD by obesity, SIRD by severe insulin resistance and MARD by mild metabolic-endocrine disturbances. In WellGen, nephropathy and retinopathy were more prevalent in SIDD compared to MOD while the latter had higher prevalence of neuropathy.

**Conclusions /Interpretation:** Our data identified insulin deficiency as the major driver of type 2 diabetes in young Indians, unlike in young European patients in whom obesity and insulin resistance predominate. Our results provide useful clues to pathophysiological mechanisms and susceptibility to complications in young Indian type 2 diabetes, and suggest a need to review management strategies.

## Introduction

Type 2 diabetes has been traditionally considered one disease characterised by both insulin resistance and insulin deficiency. Nevertheless, the disease is heterogeneous [1]. A formal description of five distinct subgroups was proposed in a large Swedish cohort [2] and replicated in other populations [2-5]. These subgroups the severely autoimmune (SAID), severely insulin-deficient (SIDD), severely insulin-resistant (SIRD), mild obesity-related (MOD) and mild age-related diabetes (MARD). The subgroups differed not only with respect to clinical characteristics at diagnosis, but also with pathophysiological mechanisms and susceptibility to complications.

India is referred to as one of the diabetes capitals of the world, and Indian type 2 diabetes patients differ from Europeans in that they develop diabetes at a younger age and are thinner [6, 7]. Indians also differ in body composition, having higher fat and lower lean proportions at the same BMI [8]. Given the role of adiposity in insulin resistance, it has therefore been assumed that type 2 diabetes in Indians is primarily driven by insulin resistance [9]. However, it is increasingly recognised that insulin deficiency may be a significant driver of diabetes in Indians [10]. Recent studies show that both the increase in the diabetes prevalence and characteristics of these patients vary in different parts of India [11, 12]. Lean type 2 diabetes is prevalent in India, especially in undernourished regions [13]. Recent studies have shown that subgroups of type 2 diabetes in Indians showed partial concordance with those in Europeans [14, 15].

Type 2 diabetes is diagnosed at a younger age in India and its pathophysiology and heterogeneity warrants further investigation. Younger age at diagnosis has distinct implications for treatment, long-term complications, mortality as well as socioeconomic burden [16]. Therefore, early identification of sub-classes may be vital for appropriate treatment to reduce adverse outcomes [17]. To address this, we implemented the Swedish algorithm [2] to identify subgroups of young Indian type 2 diabetes patients diagnosed before 45 years of age from the WellGen cohort from Pune, India [18]. We then compared Indian and European type 2 diabetes subgroups to obrain information on the relative distributions and characteristics in the two populations. We also performed *de novo* clustering of patients from the WellGen study, to assess if similar clusters are obtained sans prior hypothesis. Finally, we investigated the subclassification of type 2 diabetes in cohorts from two other geographical regions across India.

## Methods

### Study population

#### WellGen (Pune, Maharashtra, Western India)

The WellGen Study includes patients visiting the Diabetes Unit, KEM Hospital, Pune and associated clinics for routine diabetes management between 2004 and 2006 [18]. Patients diagnosed with diabetes below 45 years of age using the WHO guidelines were included [1]. Diagnosis of type 2 diabetes was based on clinical criteria: age at diagnosis more than 20 years, no history of ketoacidosis, central obesity (Waist Hip Ratio > 0.80 in women and > 0.90 in men), and response to treatment with oral antidiabetic agents. Patients with clinical diagnosis of type 1 diabetes (diagnosis before 20 years of age, history of ketoacidosis, continuous insulin treatment since diagnosis), fibrocalculous pancreatic diabetes (FCPD) or fulfilling criteria for monogenic diabetes were excluded. In total, 1612 patients were included (Table 1).

**Table 1:**
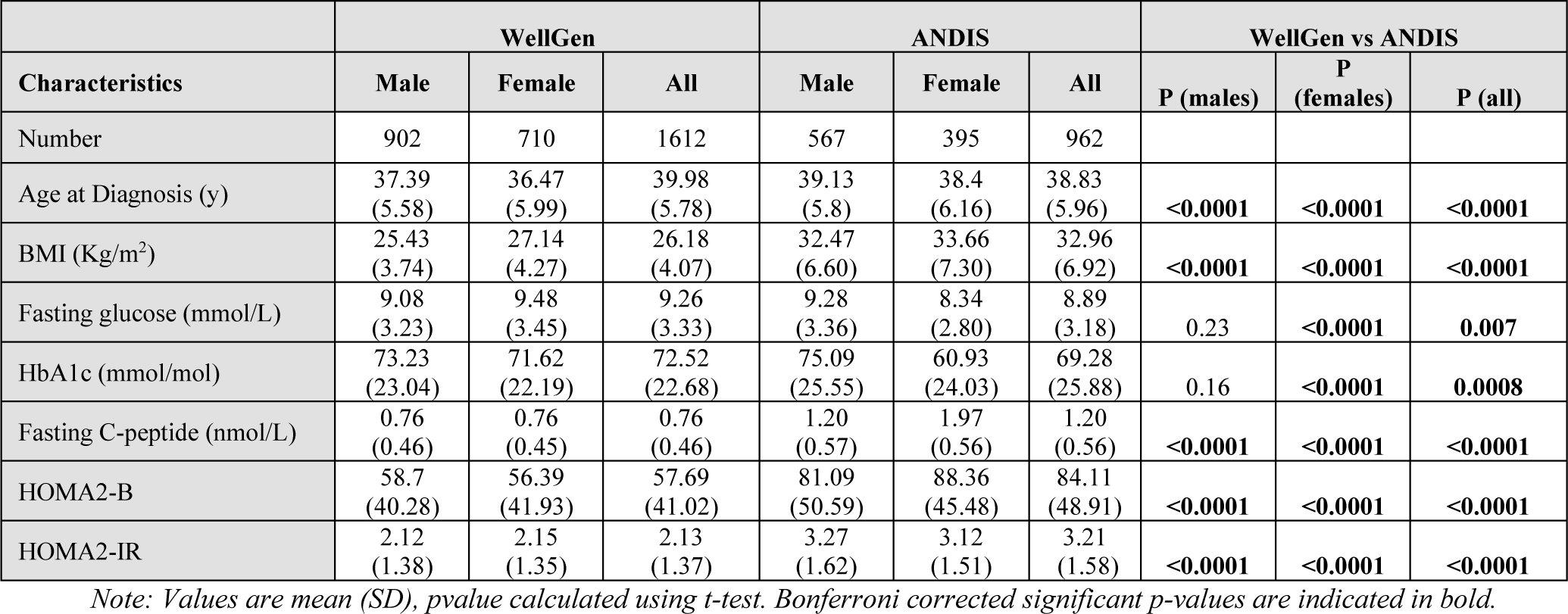
Clinical and Biochemical characteristics of participants enrolled in WellGen and ANDIS study with age at diagnosis less than 45years.

Clinical information including age, sex, age at diagnosis, family history, and socioeconomic status was obtained through a standardized questionnaire. Height, weight, waist and hip circumferences, and blood pressure were measured using standardized methods [18, 19]. Fasting plasma glucose, total and HDL cholesterol, triglycerides, and HbA1c were measured using standard laboratory assays as described earlier [18, 19]. Fasting C-peptide was measured by ELISA (Diagnostic Biochem Canade Inc, Ontario, Canada). Fasting glucose and C-peptide measurements were used to calculate HOMA2-B and HOMA2-IR values [20, 21]. Details of treatment (insulin, oral antidiabetic drugs, blood pressure and lipid-lowering medication) were recorded.

##### Complications

Coronary artery disease (CAD) was defined by ICD 10 codes I20-21, I24, I251 and I253-259. Stroke was defined by ICD 10 codes I60-61 and I63-64. Nephropathy was diagnosed by urine strip albumin measurement by a strip (nil, trace, and +), and by eGFR calculation (ml/min/1.73 m^2^ body surface area) by MDRD formula (> 90 normal, CKD: 90–60 mild, 60-30 moderate and <30 severe). Diagnosis of retinopathy was based on dilated fundus examination performed by an ophthalmologist, classified into NPDR (non-proliferative diabetic retinopathy) and PDR (proliferative diabetic retinopathy). Peripheral neuropathy was diagnosed by Biothesiometer (non-perception of vibration sense at 15 or higher amperes at two or more sites on the feet) (Table 2).

**Table 2:** Complications and current treatment by gender for participants of the WellGen and ANDIS Studies.

|  | WellGen |  |  |  | ANDIS |  |  |  |
| --- | --- | --- | --- | --- | --- | --- | --- | --- |
|  | Male (%) | Female (%) | Total (%) | p-value | Male (%) | Female (%) | Total (%) | p-value |
|  | 902 (56.0) | 710 (44.0) | 1612 |  | 567 (58.9) | 395 (41.1) | 962 |  |
| Duration of diabetes (y) | 9.73 (8.57) | 9.70 (7.74) | 9.72 (8.21) |  | 4.27 (2.56) | 4.27 (2.34) | 4.27 (2.47) |  |
| Current treatment |  |  |  |  |  |  |  |  |
| Only on Diet | 77 (8.5) | 58 (8.2) | 135 (8.4) | 0.348 | 34 (6.0) | 43 (10.9) | 77 (8.0) | **0.006** |
| Only on OHA | 558 (61.9) | 421 (59.3) | 979 (60.7) |  | 382 (67.4) | 247 (62.5) | 629 (65.4) | 0.121 |
| Only on Insulin | 51 (5.7) | 34 (4.8) | 85 (5.3) |  | 28 (4.9) | 31 (7.8) | 59 (6.1) | 0.064 |
| Both OHA+ Insulin | 216 (23.9) | 197 (27.7) | 413 (25.6) |  | 122 (21.5) | 72 (18.2) | 194 (20.2) | 0.211 |
| Complications |  |  |  |  |  |  |  |  |
| Cardiovascular disease | 106 (11.7) | 40 (4.4) | 146 (9.1) | **0.0001** | 27 (48) | 7 (1.8) | 34 (3.6) | 0.013 |
| Coronary Events | 89 (9.9) | 31 (4.4) | 120 (7.4) | **0.0001** | 19 (3.3) | 4 (1.0) | 23 (2.4) | 0.019 |
| Stroke | 17 (1.9) | 9 (1.3) | 26 (1.7) | 0.199 | 8 (1.4) | 3 (0.8) | 11 (1.2) | 0.348 |
| Nephropathy (Proteinuria and / or CKD) | 361 (40.0) | 200 (28.2) | 561 (34.8) | **0.0001** | 181 (45.9) | 129 (45.7) | 310 (45.9) | 0.960 |
| Macroalbuminuria<sup>1</sup> | 167 (19.2) | 83 (12.3) | 250 (16.2) | **0.0001** | 61 (17.1) | 29 (11.4) | 90 (14.7) | 0.047 |
| CKD: e-Glomerular Filtration Rate*<sup>2</sup> | 268 (32.3) | 145 (22.7) | 413 (28.1) | **0.0001** | 142 (26.0) | 109 (28.7) | 251 (27.1) | 0.359 |
| Early CKD (60-90) | 208 (25.0) | 122 (19.1) | 330 (22.4) | **0.0001** | 132 (24.6) | 105 (27.9) | 237 (26.0) | 0.257 |
| Moderate (30-60) | 54 (6.5) | 20 (3.1) | 74 (5.0) |  | 7 (1.3) | 4 (1.1) | 11 (1.2) | 0.747 |
| Severe (<30) | 6 (0.7) | 3 (0.5) | 9 (0.6) |  | 3 (0.5) | 0 (0.0) | 3 (0.3) | 0.148 |
| Diabetic Retinopathy (n=657)<sup>3</sup> | 102 (29.4) | 78 (25.2) | 180 (27.4) | 0.225 | 34 (19.4) | 23 (20.0) | 57 (19.7) | 0.905 |
| NPDR | 93 (26.8) | 74 (23.9) | 167 (25.4) | 0.307 | 33 (19.0) | 22 (19.3) | 55 (19.1) | 0.944 |
| PDR | 9 (2.6) | 4 (1.3) | 13 (2.0) |  | 1 (0.6) | 1 (0.9) | 2 (0.7) | 0.764 |
| Neuropathy<sup>4</sup> | 359 (40.7) | 351 (50.6) | 710 (45.1) | **0.0001** | 14 (2.5) | 2 (0.5) | 16 (1.7) | 0.019 |

Values are number (%) , \* Based on MDRD formula, p-value by Chi-square test. Bonferroni corrected significant p-values are indicated in bold.
1. Data available for n = 1020 for WellGen, n = 618 for ANDIS 2. n = 1039 for WellGen and n = 938 for ANDIS 3. n = 441 for WellGen and n = 297 for ANDIS 4. n = 1031, Neuropathy diagnosed using Biothesiometry for WellGen, ICD codes ICD 10 = E104 or E114 for ANDIS

#### Ahmedabad (Gujarat, Western India)

Patients with clinical diagnosis of type 2 diabetes visiting the DiaCare Clinic, Ahmedabad during 2018-2019 diagnosed below 45 years of age and duration of diabetes less than 2 years were invited to participate and 187 patients consented. Measurements included anthropometry, HbA1c levels, a fasting blood measurement of plasma glucose and C-peptide (MAGLUMI™ C-peptide (CLIA), Shenzhen, China). Anti-diabetic and other medication history was recorded (Supplementary table 6).

#### PHENOEINDY-2 (Dibrugarh, Assam, North-east India)

Patients with clinical diagnosis of type 2 diabetes attending medical outpatients of Assam Medical College, Dibrugarh during 2017 to 2019 if diagnosed below 40 years of age were invited to participate and 205 patients consented. Measurements included anthropometry, HbA1c levels, a fasting blood measurement of plasma glucose and C-peptide (ELISA, Diagnostic Biochem Canade Inc, Ontario, Canada). Anti-diabetic and other medication history were recorded (Supplementary table 8).

#### ANDIS (Scania, Southern Sweden)

The ANDIS project comprises newly diagnosed diabetic patients aged >18 years in the Scania County, Sweden between 2008 to 2016 [2]. Biochemical and anthropometric measurements and presence of complications were recorded as described before [2]. For the current study, 962 patients diagnosed with type 2 diabetes before 45 years of age were included. We excluded patients with known type 1 diabetes, monogenic diabetes and GAD antibody positivity (so called SAID) to maintain concordance with the WellGen study. The prevalence of complications (diagnosed as described previously [2]) was recorded on average 4.2 years after diagnosis. (Table 2).

#### DIREVA (Vaasa, Western Finland)

DIREVA includes 5107 individuals with diabetes recruited from 2009 to 2014 in the Vaasa Hospital District. For the current study, 424 type 2 diabetes patients diagnosed below 45 years of age were included, exclusion criteria were similar to ANDIS. Biochemical and anthropometric measurements have been described before [2]. No treatment or complication data from DIREVA have been included in the current study (Supplementary table 4).

All studies were approved by the local/regional Institutional Ethics Committees, and all subjects gave written informed consent.

#### Statistical methods

Patients with measurements above or below 5 standard deviations from the mean for the clustering parameters were excluded from the analysis and values outside the limits for HOMA2 calculation (fasting glucose (FG) < 3 mmol/L or FG > 25 mmol/L, or C-peptide <0.2 ng/ml or C-peptide >3.5 ng/ml) were capped to the proximal upper or lower limits. To perform supervised clustering in relation to the European derived cluster coordinates, phenotypes (age at diagnosis, HbA1c, HOMA2B, HOMA2IR and BMI) were scaled using the same scaling parameters (mean and standard deviation) as described previously [2]. Due to the unavailability of GADA data in the Indian study (WellGen), we only included clusters 2-5 (SIDD, SIRD, MOD and MARD). Patients were assigned to the pre-determined clusters on the basis of which ANDIS cluster they were most similar to, calculated as their Euclidean distance from the nearest cluster centre derived from ANDIS co-ordinates. The nearest centroid method was used to find the nearest centroid (as measured with Euclidean distances) for each patient. This resulted in each patient being assigned to any of the 4 clusters: 2/SIDD, 3/SIRD, 4/MOD or 5/MARD. Given the wide range of duration of diabetes in WellGen, we performed a sensitivity analysis by separately assessing the type 2 diabetes subgroups among those within 5-years of diagnosis and those above.

To perform unsupervised clustering, all previously mentioned variables were used in a separate analysis. Given that the results from the supervised clustering analysis showed a strong bias towards the 2/SIDD and 4/MOD cluster, and the silhouette analysis indicated that two was the most stable number of clusters, we performed k-means clustering into 2 clusters. All phenotypes were scaled to have a mean of 0 and a standard deviation of 1, this time with scaling parameters derived from the data itself. k-means clustering was then performed separately in females and males using the k-means runs algorithm from the fpc R package.

#### Type 1 diabetes risk scores

In absence of GADA data, we applied validated ‘type 1’ Genetic Risk Score (GRS) to 560 WellGen participants with available data to estimate the proportion of those carrying autoimmune risk alleles. These patients did not differ from those on whom genotyping was not available (ESM table 5). Positive controls were 261 type 1 diabetic patients as described previously [22]. Negative controls were 461 normal glucose tolerant (75 g oral glucose tolerance test; WHO 1999 criteria) participants from the Pune Maternal Nutrition Study [23].

##### Genotyping

Genome wide genotyping data was generated on WellGen and PMNS participants using Affymetrix SNP 6.0 Chips (Affymetrix, CA, USA) and the Infinium Global Screening Array V1 B37 (Illumina) for type 1 diabetes cohort. QC and Imputation were performed as described in ESM methods. In addition, Sequenom Mass Array technology was used to validate the 9 type 1 diabetes associated SNPs (Supplementary Table 5) with >98% success rate in the type 1 diabetes cohort.

#### GRS

A previously described set of 9 SNPs was used for type 1 diabetes GRS calculations (Supplementary Table 5) [22, 24, 25]. In the absence of genotyping data for rs7454108, a proxy SNP rs3957146 (LD: r2=1, D’=1) was used. The haplotype was constructed using rs2187668+ rs3957146 as described previously [22] and GRS scores were computed on plink using weighted scores. Logistic regression was performed to assess the discriminatory power of GRS between type 1 diabetes and other subgroups.

## Results

We first sought to investigate the subgroups of young type 2 diabetes patients in Indian WellGen study and compare them with Swedish subgroups from the ANDIS study.

The WellGen study comprised patients diagnosed with type 2 diabetes before 45 years of age, and all relevant data required for clustering were available on 1624 patients. After applying exclusions, 1612 patients (56% male) with average age at diagnosis of 40 years, duration of diabetes ∼10 years, and BMI 26.18 kg/m^2^ were included. (ESM fig 1, table 1).

**Figure 1.**
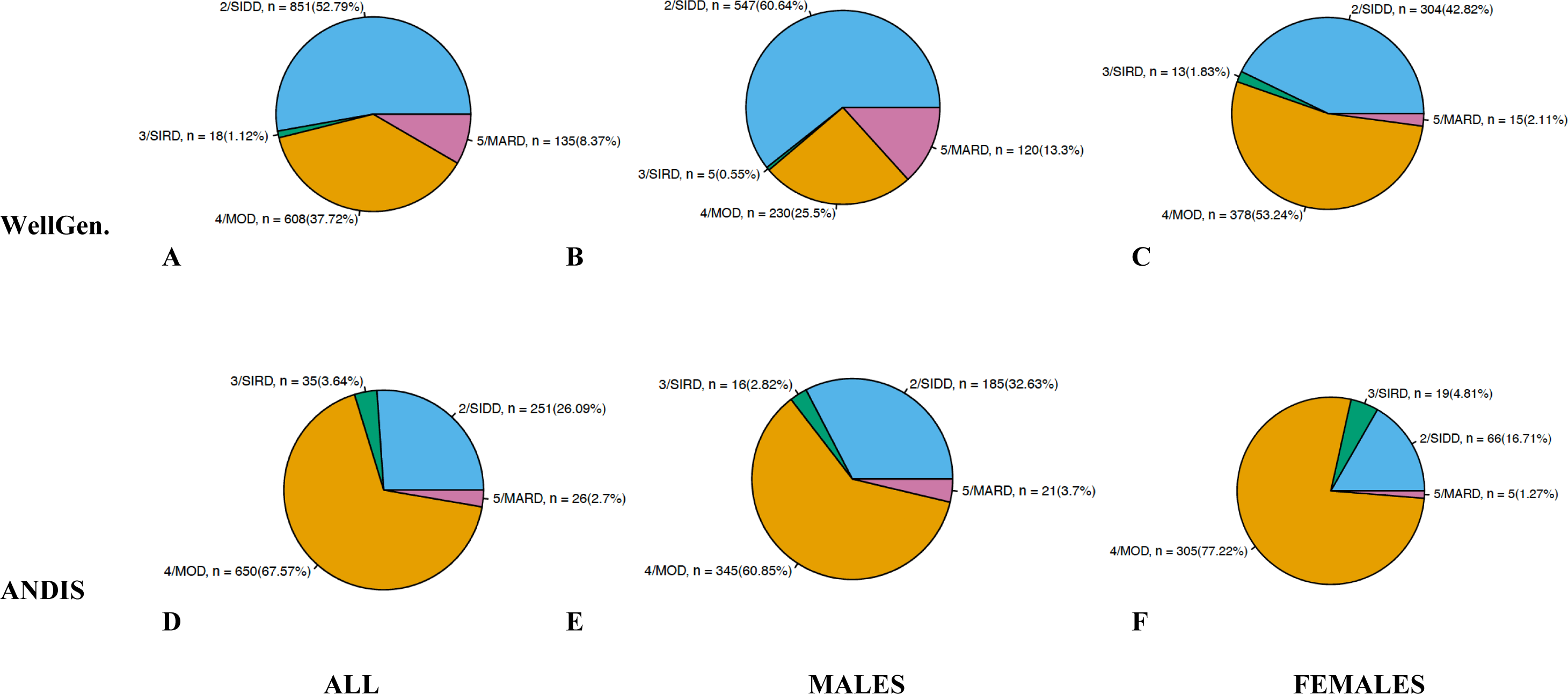
Distribution of participants from the WellGen and ANDIS study in the predefined clusters. (A) Distribution of WellGen patients (n = 1612) (B) distribution of men with diabetes from the WellGen study (n = 902) (C) distribution of women with diabetes from the WellGen study (n =710) (D) Distribution of ANDIS patients (n = 973) (E) distribution of men with diabetes from the ANDIS study (n = 575) (F) distribution of women with diabetes from the ANDIS study (n = 398)

For comparison, we selected 962 type 2 diabetes patients (58.9% male, average age at diagnosis 38.83 years) from the ANDIS study, after excluding 577 patients belonging to cluster 1 (SAID) [2]. The Indian patients were younger at diagnosis, had lower BMI, higher fasting plasma glucose, and lower fasting C-peptide, HOMA2B and HOMA2IR compared to the ANDIS sub-cohort (Table 1). The proportion of patients on lifestyle management alone, on antidiabetic oral agents and on insulin treatment was broadly similar in both cohorts (Table 2). In the WellGen study, men had a higher prevalence of cardiovascular events, nephropathy and retinopathy compared to women whereas that of neuropathy was higher in women. There was no difference with respect to these complications between men and women in the ANDIS study (Table 2). We did not compare the complication rates between the two cohorts because of difference in duration of diabetes.

### SIDD predominant in India, MOD in Sweden

In the absence of GADA data, we obtained the four expected clusters, albeit with different proportions. In the WellGen study, the severe insulin-deficient SIDD cluster was the largest subgroup (52.8%), followed by the mild obesity-related MOD (37.7%), while severely insulin resistant SIRD (1.1%) and mild age-related MARD (8.4%) were less common. In a sensitivity analysis, with increasing duration of diabetes (from less than to more than 5y), the proportion of patients in the SIDD subgroup increased (45.5 to 56.9%) while that in MOD decreased (44.7 to 33.8%) (ESM Table 1).

In the sex-stratified analysis, SIDD (60.6%) remained the predominant cluster in the men whereas MOD (53.2%) in women; MARD was more common in men (13.3% vs 2.1%) (Figure 1, Table 3).

**Table 3:** Characteristics of participants enrolled in WellGen and ANDIS study by clusters for all participants, males and females.

|  | WellGen |  |  |  |  |  | ANDIS |  |  |  |  |  | WellGen vs ANDIS |  |
| --- | --- | --- | --- | --- | --- | --- | --- | --- | --- | --- | --- | --- | --- | --- |
| Gender | All (1612) |  |  |  |  |  | All (962) |  |  |  |  |  |  |  |
| Cluster | SIDD | SIRD | MOD | MARD | p | p1 | SIDD | SIRD | MOD | MARD | p | p1 | p: SIDD | p:MOD |
| Number | 851<br>(52.79) | 18<br>(1.11) | 608<br>(37.71) | 135<br>(8.37) |  |  | 251<br>(26.09) | 35<br>(3.64) | 650<br>(67.56) | 26<br>(2.7) |  |  |  |  |
| Age at Diagnosis (y) | 36.86<br>(5.81) | 38.83<br>(4.93) | 36.25<br>(5.77) | 40.84<br>(3.98) | 0.005 | 0.0001 | 38.98<br>(6.24) | 39.94<br>(5.87) | 38.56<br>(5.9) | 42.5<br>(2.79) | < 0.0001 | < 0.0001 | < 0.0001 | < 0.0001 |
| Duration of diabetes (y) | 10.49<br>(8.25) | 6.41<br>(7.16) | 8.72<br>(8.03) | 9.83<br>(8.44) | 0.001 | -- | -- | -- | -- | -- | -- | -- | -- | -- |
| BMI (Kg/m<sup>2</sup>) | 25.02<br>(3.45) | 28.37<br>(5.27) | 28.4<br>(4.03) | 23.22<br>(2.44) | 0.0001 | 0.0001 | 28.29<br>(5.43) | 36.62<br>(6.28) | 34.92<br>(6.41) | 24.11<br>(2.33) | < 0.0001 | < 0.0001 | <0.0001 | < 0.0001 |
| Fasting glucose ( mmol/L) | 10.77<br>(3.39) | 6.86<br>(1.89) | 7.85<br>(2.37) | 6.50<br>(1.47) | 0.0001 | 0.0001 | 11.75<br>(3.67) | 7.05<br>(2.22) | 7.97<br>(2.25) | 6.75<br>(1.26) | < 0.0001 | < 0.0001 | <0.0001 | 0.06 |
| HbA1c (mmol/mol) | 87.01<br>(19.34) | 56.7<br>(13.1) | 58.33<br>(12.92) | 47.25<br>(10.37) | 0.0001 | 0.0001 | 100.6<br>(19.62) | 55.19<br>(13.79) | 58.87<br>(17.64) | 45.85<br>(7.41) | < 0.0001 | < 0.0001 | < 0.0001 | 0.53 |
| Fasting C-Peptide (nmol/L) | 0.68<br>(0.39) | 1.01<br>(0.62) | 0.89<br>(0.48) | 0.58<br>(0.29) | 0.0001 | 0.0001 | 0.85<br>(0.4) | 2.53<br>(0.61) | 1.29<br>(0.48) | 0.73<br>(0.24) | < 0.0001 | < 0.0001 | < 0.0001 | <0.0001 |
| HOMA2B | 37.66<br>(22.24) | 214.99<br>(41.01) | 77.49<br>(42.02) | 73.74<br>(31.16) | 0.0001 | 0.0001 | 41.03<br>(28.34) | 968.71<br>(56.43) | 94.79<br>(38.53) | 80.15<br>(27.62) | < 0.0001 | < 0.0001 | 0.048 | <0.0001 |
| HOMA2IR | 2.08<br>(1.41) | 4.73<br>(1.56) | 2.28<br>(1.29) | 1.42<br>(0.76) | 0.314 | 0.0001 | 2.67<br>(1.42) | 6.17<br>(1.86) | 3.31<br>(1.42) | 1.78<br>(0.63) | < 0.0001 | < 0.0001 | 0.043 | <0.0001 |

| Gender | Male (902) |  |  |  |  |  | Male (567) |  |  |  |  |  |  |  |
| --- | --- | --- | --- | --- | --- | --- | --- | --- | --- | --- | --- | --- | --- | --- |
| Cluster | SIDD | SIRD | MOD | MARD | p | p1 | SIDD | SIRD | MOD | MARD | p | p1 | p: SIDD | p:MOD |
| Number | 547<br>(60.6) | 5<br>(0.55) | 230<br>(25.49) | 120<br>(13.30) |  |  | 185<br>(32.6) | 16<br>(2.82) | 345<br>(60.84) | 21<br>(3.7) |  |  |  |  |
| Age at Diagnosis (y) | 36.96<br>(5.67) | 42.07<br>(10.27) | 36.59<br>(5.43) | 40.7<br>(4.12) | 0.0001 | 0.0001 | 38.91<br>(6.08) | 40.56<br>(6.19) | 39.0<br>(5.73) | 42.05<br>(2.87) | 0.08 | 0.089 | <0.0001 | <0.0001 |
| Duration of diabetes (y) | 10.55<br>(8.70) | 8.03<br>(6.59) | 7.71<br>(7.98) | 9.98<br>(8.56) | 0.005 | -- |  |  |  |  |  |  |  |  |
| BMI (Kg/m<sup>2</sup>) | 24.44<br>(3.08) | 28.75<br>(3.54) | 28.77<br>(3.67) | 23.41<br>(2.34) | 0.0001 | 0.0001 | 27.45<br>(3.9) | 36.74<br>(6.01) | 35.44<br>(5.9) | 24.72<br>(2.11) | <0.0001 | <0.0001 | <0.0001 | <0.0001 |
| Fasting glucose ( mmol/L) | 10.29<br>(3.22) | 5.75<br>(1.0) | 7.63<br>(2.43) | 6.52<br>(1.53) | 0.0001 | 0.0001 | 11.80<br>(3.5) | 7.58<br>(3.04) | 8.16<br>(2.53) | 6.77<br>(2.53) | <0.0001 | <0.0001 | <0.0001 | 0.013 |
| HbA1c (mmol/mol) | 84.67<br>(20.57) | 49.07<br>(10.27) | 59.94<br>(13.35) | 47.55<br>(10.02) | 0.0001 | 0.0001 | 99.58<br>(20.31) | 57.33<br>(13.82) | 64.59<br>(18.46) | 45.30<br>(7.11) | <0.0001 | <0.0001 | <0.0001 | 0.001 |
| Fasting C-Peptide (nmol/L) | 0.66<br>(0.38) | 2.17<br>(0.44) | 1.08<br>(0.51) | 0.60<br>(0.30) | 0.0001 | 0.0001 | 0.82<br>(0.35) | 2.65<br>(0.61) | 1.36<br>(0.49) | 0.72<br>(0.49) | <0.0001 | <0.0001 | <0.0001 | <0.0001 |
| HOMA2B | 39.33<br>(22.5) | 229.16<br>(40.98) | 92.27<br>(42.49) | 75.58<br>(31.92) | 0.0001 | 0.0001 | 39.15<br>(25.31) | 200.56<br>(74.37) | 98.19<br>(40.98) | 78.57<br>(27.41) | <0.0001 | <0.0001 | 0.92 | 0.094 |
| HOMA2IR | 1.96<br>(1.34) | 4.99<br>(1.23) | 2.76<br>(1.41) | 1.47<br>(0.78) | 0.092 | 0.0001 | 2.59<br>(1.27) | 6.67<br>(2.23) | 3.56<br>(1.48) | 1.77 (0.65) | <0.0001 | <0.0001 | <0.0001 | <0.0001 |

| Gender | Female (710) |  |  |  |  |  | Female (398) |  |  |  |  |  |  |  |
| --- | --- | --- | --- | --- | --- | --- | --- | --- | --- | --- | --- | --- | --- | --- |
| Cluster | SIDD | SIRD | MOD | MARD | p | p1 | SIDD | SIRD | MOD | MARD | p | p1 | p: SIDD | p:MOD |
| Number | 304<br>(42.81) | 13<br>(1.83) | 378<br>(53.23) | 15<br>(2.11) |  |  | 66<br>(16.7) | 19<br>(4.8) | 305<br>(77.2) | 5<br>(1.26) |  |  |  |  |
| Age at Diagnosis (y) | 36.68<br>(6.06) | 37.51<br>(4.92) | 36.05<br>(5.96) | 41.98<br>(2.51) | 0.722 | 0.001 | 39.2<br>(6.72) | 39.42<br>(5.71) | 38.07<br>(6.06) | 44.4<br>(1.34) | 0.03 | 0.045 | **0.0028** | **<0.0001** |
| Duration of diabetes (y) | 10.38<br>(7.37) | 5.79<br>(7.53) | 9.34<br>(8.00) | 8.57<br>(7.53) | 0.070 | -- |  |  |  |  |  |  |  |  |
| BMI (Kg/m<sup>2</sup>) | 26.07<br>(3.82) | 28.23<br>(5.92) | 28.18<br>(4.22) | 21.69<br>(2.75) | 0.0001 | 0.0001 | 30.65<br>(7.92) | 36.51<br>(6.67) | 34.33<br>(6.90) | 21.54<br>(1.19) | **<0.0001** | **<0.0001** | **<0.0001** | **<0.0001** |
| Fasting glucose ( mmol/L) | 11.64<br>(3.53) | 5.89<br>(1.44) | 7.98<br>(2.33) | 6.33<br>(0.97) | 0.0001 | 0.0001 | 11.61<br>(4.11) | 6.63<br>(1.08) | 7.76<br>(1.88) | 6.66<br>(1.62) | **<0.0001** | **<0.0001** | 0.92 | 0.19 |
| HbA1c (mmol/mol) | 91.22<br>(16.11) | 59.63<br>(13.21) | 57.34<br>(12.57) | 44.85<br>(12.98) | 0.0001 | 0.0001 | 103.48<br>(17.35) | 53.39<br>(13.88) | 52.41<br>(14.12) | 48.15<br>(9.04) | **<0.0001** | **<0.0001** | **<0.0001** | **<0.0001** |
| Fasting C-Peptide (nmol/L) | 0.71<br>(0.41) | 1.98<br>(0.63) | 0.77<br>(0.42) | 0.42<br>(0.20) | 0.479 | 0.0001 | 0.90<br>(0.5) | 2.43<br>(0.61) | 1.19<br>(0.46) | 0.76<br>(0.23) | **<0.0001** | **<0.0001** | **0.0013** | **<0.0001** |
| HOMA2B | 34.67<br>(21.47) | 209.54<br>(41.32) | 68.5<br>(39.13) | 59.05<br>(19.27) | 0.0001 | 0.0001 | 46.29<br>(35.19) | 193.47<br>(37.16) | 90.94<br>(35.22) | 86.78<br>(30.71) | **<0.0001** | **<0.0001** | **0.00052** | **<0.0001** |
| HOMA2IR | 2.29<br>(1.49) | 4.63<br>(1.71) | 1.98<br>(1.12) | 1.03<br>(0.47) | 0.0001 | 0.0001 | 2.9<br>(1.78) | 5.74<br>(1.39) | 3.03<br>(1.30) | 1.84<br>(0.61) | **<0.0001** | **<0.0001** | **0.0043** | **<0.0001** |

Note: Values are mean (SD), p-value by ANOVA, p1 adjusted for duration of diabetes. Bonferroni corrected significant p-values are indicated in bold.

Concordant to the diabetes subgroups in ANDIS, Indian SIDD patients had the lowest insulin secretion (HOMA-ß) and the highest glycemia while MOD had the highest BMI. SIRD were the most insulin resistant, whereas MARD were the oldest at diagnosis, with mildest glycemia and lowest degree of insulin resistance (Figure 2, Table 3). These results support the pathophysiological basis for subclassification in a population which has a different genetic and socio-economic background compared to the Swedish.

**Figure 2.**
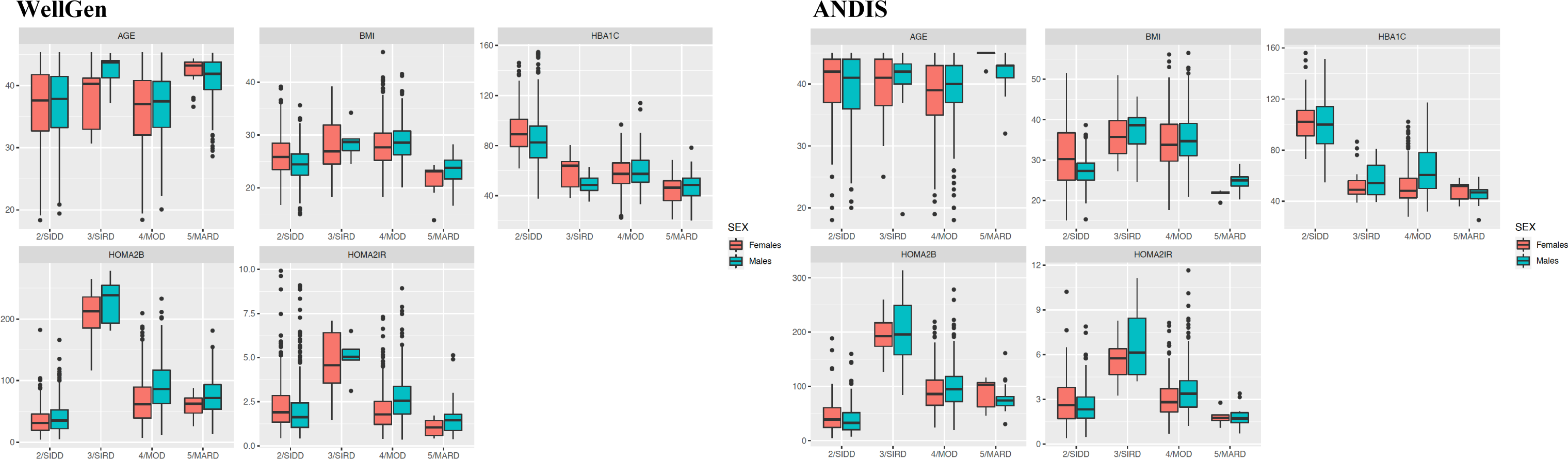
Cluster Characteristics in the WellGen and ANDIS studies. Distribution of age at diagnosis, BMI, HbA1c, HOMA2-B and HOMA2-IR in the WellGen and ANDIS studies for each cluster. k-means clustering was done separately for men and women; data is shown for each sex separately. SIDD = severe insulin-deficient diabetes, SIRD = resistant diabetes, MOD = mild obesity-related-related diabetes, MARD = mild age-related diabetes. HOMA2-B = homeostatic model assessment 2 estimates of beta cell function. HOMA2-B = homeostatic model assessment 2 estimates insulin resistance.

The distribution of subgroups in ANDIS differed from that of WellGen; MOD was the most predominant cluster (67.56%), followed by SIDD (26.09%), SIRD (3.64%) and MARD (2.70%). These distributions were similar in men and women (Figure 1, Table 3). The pathophysiological characteristics in these young Swedish type 2 diabetes patients were similar to those in the parent ANDIS cohort (Figure 2, Table 3).

#### Treatment

In both WellGen and ANDIS, insulin treatment (alone or in combination with OHAs) was most commonly prescribed to SIDD patients (38.3% in WellGen, 39.5% in ANDIS). (Table 4).

**Table 4:**
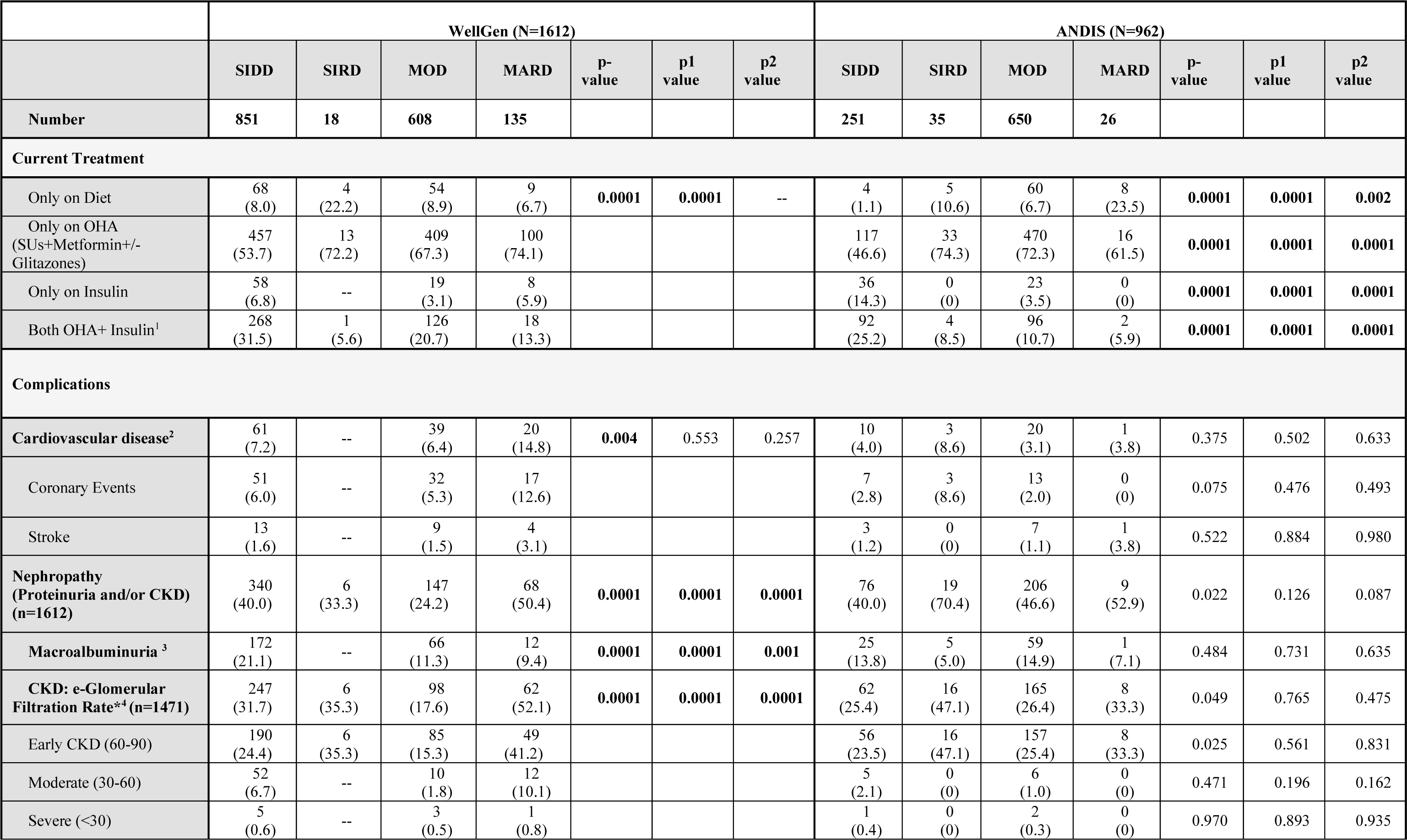
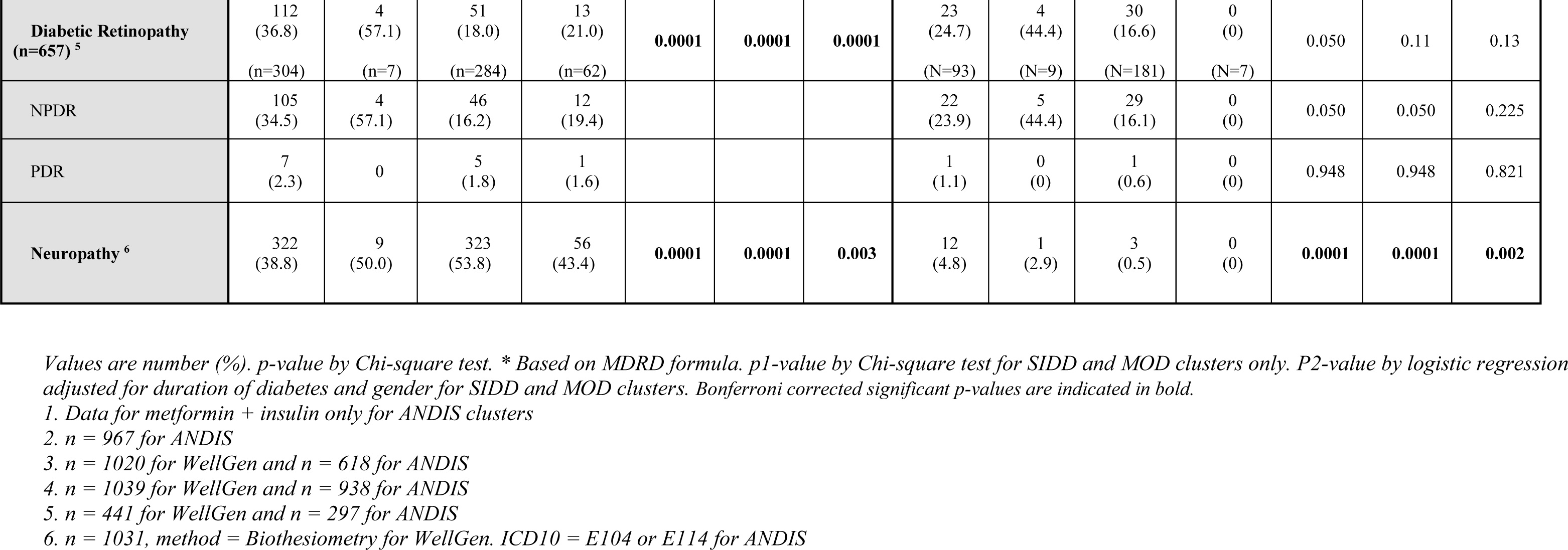
Treatment and complications by cluster in the WellGen and ANDIS study.

|  | WellGen (N=1612) |  |  |  |  |  |  | ANDIS (N=962) |  |  |  |  |  |  |
| --- | --- | --- | --- | --- | --- | --- | --- | --- | --- | --- | --- | --- | --- | --- |
|  | SIDD | SIRD | MOD | MARD | p-value | p1 value | p2 value | SIDD | SIRD | MOD | MARD | p-value | p1 value | p2 value |
| Number | 851 | 18 | 608 | 135 |  |  |  | 251 | 35 | 650 | 26 |  |  |  |
| Current Treatment |  |  |  |  |  |  |  |  |  |  |  |  |  |  |
| Only on Diet | 68<br>(8.0) | 4<br>(22.2) | 54<br>(8.9) | 9<br>(6.7) | 0.0001 | 0.0001 | -- | 4<br>(1.1) | 5<br>(10.6) | 60<br>(6.7) | 8<br>(23.5) | 0.0001 | 0.0001 | 0.002 |
| Only on OHA<br>(SUs+Metformin+/-<br>Glitazones) | 457<br>(53.7) | 13<br>(72.2) | 409<br>(67.3) | 100<br>(74.1) |  |  |  | 117<br>(46.6) | 33<br>(74.3) | 470<br>(72.3) | 16<br>(61.5) | 0.0001 | 0.0001 | 0.0001 |
| Only on Insulin | 58<br>(6.8) | -- | 19<br>(3.1) | 8<br>(5.9) |  |  |  | 36<br>(14.3) | 0<br>(0) | 23<br>(3.5) | 0<br>(0) | 0.0001 | 0.0001 | 0.0001 |
| Both OHA+ Insulin<sup>1</sup> | 268<br>(31.5) | 1<br>(5.6) | 126<br>(20.7) | 18<br>(13.3) |  |  |  | 92<br>(25.2) | 4<br>(8.5) | 96<br>(10.7) | 2<br>(5.9) | 0.0001 | 0.0001 | 0.0001 |
| Complications |  |  |  |  |  |  |  |  |  |  |  |  |  |  |
| Cardiovascular disease<sup>2</sup> | 61<br>(7.2) | -- | 39<br>(6.4) | 20<br>(14.8) | 0.004 | 0.553 | 0.257 | 10<br>(4.0) | 3<br>(8.6) | 20<br>(3.1) | 1<br>(3.8) | 0.375 | 0.502 | 0.633 |
| Coronary Events | 51<br>(6.0) | -- | 32<br>(5.3) | 17<br>(12.6) |  |  |  | 7<br>(2.8) | 3<br>(8.6) | 13<br>(2.0) | 0<br>(0) | 0.075 | 0.476 | 0.493 |
| Stroke | 13<br>(1.6) | -- | 9<br>(1.5) | 4<br>(3.1) |  |  |  | 3<br>(1.2) | 0<br>(0) | 7<br>(1.1) | 1<br>(3.8) | 0.522 | 0.884 | 0.980 |
| Nephropathy<br>(Proteinuria and/or CKD)<br>(n=1612) | 340<br>(40.0) | 6<br>(33.3) | 147<br>(24.2) | 68<br>(50.4) | 0.0001 | 0.0001 | 0.0001 | 76<br>(40.0) | 19<br>(70.4) | 206<br>(46.6) | 9<br>(52.9) | 0.022 | 0.126 | 0.087 |
| Macroalbuminuria<sup>3</sup> | 172<br>(21.1) | -- | 66<br>(11.3) | 12<br>(9.4) | 0.0001 | 0.0001 | 0.001 | 25<br>(13.8) | 5<br>(5.0) | 59<br>(14.9) | 1<br>(7.1) | 0.484 | 0.731 | 0.635 |
| CKD: e-Glomerular<br>Filtration Rate*<sup>4</sup> (n=1471) | 247<br>(31.7) | 6<br>(35.3) | 98<br>(17.6) | 62<br>(52.1) | 0.0001 | 0.0001 | 0.0001 | 62<br>(25.4) | 16<br>(47.1) | 165<br>(26.4) | 8<br>(33.3) | 0.049 | 0.765 | 0.475 |
| Early CKD (60-90) | 190<br>(24.4) | 6<br>(35.3) | 85<br>(15.3) | 49<br>(41.2) |  |  |  | 56<br>(23.5) | 16<br>(47.1) | 157<br>(25.4) | 8<br>(33.3) | 0.025 | 0.561 | 0.831 |
| Moderate (30-60) | 52<br>(6.7) | -- | 10<br>(1.8) | 12<br>(10.1) |  |  |  | 5<br>(2.1) | 0<br>(0) | 6<br>(1.0) | 0<br>(0) | 0.471 | 0.196 | 0.162 |
| Severe (<30) | 5<br>(0.6) | -- | 3<br>(0.5) | 1<br>(0.8) |  |  |  | 1<br>(0.4) | 0<br>(0) | 2<br>(0.3) | 0<br>(0) | 0.970 | 0.893 | 0.935 |
| **Diabetic Retinopathy**<br>**(n=657) <sup>5</sup>** | 112<br>(36.8)<br>(n=304) | 4<br>(57.1)<br>(n=7) | 51<br>(18.0)<br>(n=284) | 13<br>(21.0)<br>(n=62) | **0.0001** | **0.0001** | **0.0001** | 23<br>(24.7)<br>(N=93) | 4<br>(44.4)<br>(N=9) | 30<br>(16.6)<br>(N=181) | 0<br>(0)<br>(N=7) | 0.050 | 0.11 | 0.13 |
| NPDR | 105<br>(34.5) | 4<br>(57.1) | 46<br>(16.2) | 12<br>(19.4) |  |  |  | 22<br>(23.9) | 5<br>(44.4) | 29<br>(16.1) | 0<br>(0) | 0.050 | 0.050 | 0.225 |
| PDR | 7<br>(2.3) | 0 | 5<br>(1.8) | 1<br>(1.6) |  |  |  | 1<br>(1.1) | 0<br>(0) | 1<br>(0.6) | 0<br>(0) | 0.948 | 0.948 | 0.821 |
| **Neuropathy <sup>6</sup>** | 322<br>(38.8) | 9<br>(50.0) | 323<br>(53.8) | 56<br>(43.4) | **0.0001** | **0.0001** | **0.003** | 12<br>(4.8) | 1<br>(2.9) | 3<br>(0.5) | 0<br>(0) | **0.0001** | **0.0001** | **0.002** |

Values are number (%). p-value by Chi-square test. \* Based on MDRD formula. p1-value by Chi-square test for SIDD and MOD clusters only. P2-value by logistic regression adjusted for duration of diabetes and gender for SIDD and MOD clusters. Bonferroni corrected significant p-values are indicated in bold.
1. Data for metformin + insulin only for ANDIS clusters
2. n = 967 for ANDIS
3. n = 1020 for WellGen and n = 618 for ANDIS
4. n = 1039 for WellGen and n = 938 for ANDIS
5. n = 441 for WellGen and n = 297 for ANDIS
6. n = 1031, method = Biothesiometry for WellGen. ICD10 = E104 or E114 for ANDIS

#### Complications

In WellGen, we compared the prevalence of complications in the two major subtypes, SIDD and MOD. Small numbers in SIRD and MARD preclude comparison of complications in these subgroups. Retinopathy and nephropathy were most common in SIDD whereas neuropathy was more prevalent in MOD. Prevalence of macrovascular complications was similar in the two subtypes. Of the less common subgroups, SIRD had a high prevalence of retinopathy while MARD of nephropathy and macrovascular disease. (Table 4).

In the young ANDIS cohort, nephropathy (70.4%) and retinopathy (44.4%) prevalence were highest in SIRD whereas neuropathy was most common in SIDD. Consistent with previous findings, SIRD also showed the highest prevalence of CKD (47.1%) while macroalbuminuria (14.9%) was most common in MOD (Table 4).

### De novo clusters shows high degree of concordance with SIDD and MOD

We also applied the *de novo* k-means clustering to assess the subgroups obtained in the Indian patients and compared them with those obtained using the previously published algorithm. Two was the optimum number of clusters (ESM Fig 2a); cluster 1 had a prevalence of 66.6% while cluster 2 had a prevalence of 33.4% (ESM table 2). Cluster 1 showed 88.8% concordance with SIDD (86.8% in males, 92.4% in females) while cluster 2 had an overlap of 62.5% with MOD (83.9% in males, ∼49.5% in females) (ESM Fig 2b).

Both clusters had the same cluster characteristics as seen using the centroid method, thereby providing a technical replication (ESM Table 2). The similarity also extended to complications rates, with nephropathy and retinopathy being prevalent in cluster 1 compared to cluster 2 whereas neuropathy was more prevalent in cluster 2 (ESM Table 3).

### Low prevalence of genetic type 1 diabetes in WellGen type 2 diabetes subgroups

In the absence of GADA data, a previously established type 1 diabetes GRS [24] comprising 9 SNPs (ESM Table 4) which was validated in the Indian population [22] was applied to estimate the proportion of patients with autoimmune diabetes in a subset of the WellGen cohort (ESM Table 5). The GRS was associated with the positive control type 1 diabetic patients compared to SIDD and MOD. The same GRS did not associate with either SIDD or MOD compared to non-diabetic controls (ESM Table 6 and ESM Fig 3). The proportion of patients with GRS ≥ 90% and 80% scores was ∼ 5% and 28.7%, respectively, in type 1 diabetes (positive controls) whereas 0% and 1.4% in SIDD, 0% and 4.7% in MOD, 0% and 1% in controls. The same GRS was associated with SAID in young ANDIS patients (beta=7.3±0.72, p < 2×10^−16^).

### Indian diabetes subgroups similar to subgroups in European cohort DIREVA with longer diabetes duration

Given the difference in duration of diabetes in ANDIS and WellGen, we compared the WellGen subgroups with those in the Finnish cohort DIREVA (n=420) with duration of diabetes 14.4 years (ESM Table 7). The differences in proportion of subgroups and cluster characteristics between WellGen and DIREVA were similar to those as between WellGen and ANDIS (ESM Table 8, ESM Fig 4).

### Subgroups of diabetes in other regions of India

We applied the Swedish algorithm [2] to two studies from different geographical regions in India, Ahmedabad, Gujarat, Western India (N=187) and Dibrugarh, Assam, North-Eastern India (N=205) (ESM Table 9 - 12). Concordant to findings in WellGen, the Ahmedabad cohort had the highest proportion of SIDD patients (56.68%) followed by MOD (33.15%) and MARD (10.16%). The similarity extended to the subgroup distributions in the two sexes: The proportion of SIDD was highest in males (61.59%) while MOD in females (53.06%) (ESM Table 10, ESM Fig 5). The PHENOEINDY-2 cohort was the youngest and the thinnest cohort of all, the proportion of SIDD (66.66%) was the highest, followed by MOD (23.20%), MARD (7.72%) and SIRD (1.40%) (ESM Table 12, ESM Fig 6).

## Discussion

We showed that the clusters described in the newly diagnosed unselected European type 2 diabetes patients [2, 3] are also seen in the younger and thinner Indians. SIDD and MOD were the two predominant subgroups, while MARD was less common and SIRD the least common in both populations. SIDD was the predominant cluster in Indians whereas MOD in Europeans. The predominance of SIDD was replicated in two independent, geographically distinct Indian cohorts of patients with young onset type 2 diabetes.

The distribution of the clusters suggests that deficient insulin secretion, rather than the often-purported insulin resistance, is the driver of young type 2 diabetes in India. In contrast, in the young Swedish and Finnish type 2 diabetes patients, obesity and insulin resistance seemed to be the primary pathophysiological drivers. The proposed prominent role of insulin resistance was based on previous demonstrations of higher insulin resistance in Indians compared to Europeans at a given BMI possibly due to relatively more adipose body composition [7, 26, 27]. Despite the differences in age, BMI and duration of diabetes, the characteristics of the clusters themselves in Indians broadly reflected those in the European studies [2] Our new findings suggest a paradigm shift for the understanding of the pathophysiology of type 2 diabetes in young Indians, albeit they do not preclude the role of insulin resistance.

*De novo* k-means subclassification validated the two major subgroups obtained from the European derived centroids. The concordance was greater in the men for both subgroups, while, it was lower in women for the newly obtained cluster 2 with MOD. While, this increases our confidence in the classification, reclassification of a proportion of MOD women to a SIDD equivalent cluster 1 highlights the role of insulin deficiency in the pathogenesis of type 2 diabetes in young Indians.

We applied the European derived centroids to two smaller cohorts of young type 2 diabetes patients from western (Ahmedabad, Gujarat) and north-eastern (Dibrugarh, Assam) India. The proportion of subclasses in Ahmedabad patients was nearly identical to those in Pune while the proportion of SIDD patients was highest in Dibrugarh. Gujarat is a more affluent state while Assam has a lower development index and high prevalence of undernutrition.

Physicians in India have long realized the phenotypic differences of Indian diabetes patients compared to those described in patients of European origin [28, 29]. Interestingly, the lean type 2 diabetes has been prominently reported from impoverished states of Orissa and north-east India, where malnutrition related diabetes (MRDM) was described [30]. A proportion of SIDD patients from Assam could well be characterised similarly. There is an increasing recognition that early life undernutrition could lead to smaller beta-cell mass and insulin secretion defects demonstrable from early childhood in serially studied birth cohorts and manifest as pre-diabetes or type 2 diabetes in young adulthood [31, 32]. Animal studies have clearly demonstrated poor beta-cell development and islet dysfunction in the offspring born to malnourished pregnant mothers [33-35]. It is intriguing that the highest rise in the prevalence of type 2 diabetes in India over the last 25 years has been demonstrated in states which have suffered chronic environmental, socioeconomic and nutritional deficits [36]. On such a background of intergenerational deprivation, a relatively small socioeconomic development appears enough to precipitate diabetes at young age. It is of note that the prevalence of diabetes in those above 20 years of age has increased from 5.5 to 7.5% between 1990 and 2016 in the state of Assam.

The subgroups had different sensitivities to micro- and macrovascular complications. Microvascular affection in retina and kidney was more prevalent in SIDD compared to MOD while peripheral nerve affection was more prevalent in MOD. Possible reasons for these differences may lie in the pathophysiological mechanisms driving the subgroups which need to be studied further. Macrovascular disease was similar in two subgroups. In the original Swedish classification, SIRD subgroup generated lot of interest given their high propensity to develop nephropathy. It was the smallest subgroup in Indians with high insulin resistance as well as insulin secretion, however, was heterogeneous between the Indian cohorts. Intriguingly, MARD subgroup had a strikingly high rate of Macrovascular disease. The unique profiles of these subgroups could well represent population-specific differences and highlight the need for customisation of the clustering algorithm.

Other studies have investigated the heterogeneity of type 2 diabetes in Indians: The INSPIRED study from a chain of private diabetes clinics in India reported four subclasses, two of which were similar to the Swedish study (SIDD and MARD) [5], and two were new (insulin resistant obese diabetes – IROD and combined insulin resistance and deficient diabetes - CIRDD) [14]. However, the clustering parameters were different and therefore not directly comparable with our study. The MASALA-MESA study reported subclasses in a mixed population in the USA including migrant South Asian Indians (n=217). They found an excess of younger, thinner and severely hyperglycemic patients in the South Asian Indians supporting our findings [15].

Strengths and limitations: This is the first attempt in India to subclassify patients with a diagnosis of type 2 diabetes at a young age. The presence of comparable subgroups in our study to a genetically and historically distinct European population validates the subgroups. While the power of the study is limited, validation by *de novo* clustering increases confidence in classification. The Indian patients are clinic-based and enrolled many years after diagnosis while on antidiabetic treatment which may affect the proportions of subclasses. We cannot rule out the possibility that some patients might shift to different subgroups over time, but only a small proportion did so in another study [3]. A sensitivity analysis in WellGen showed that the proportions varied with increasing duration, however, SIDD (45.5%) remained the predominant subgroup even in those with less than 5 years of diagnosis. Observations in other two Indian cohorts further validated these findings. Another limitation is that this study is an opportunistic comparison of existing data and therefore laboratory measurements are not fully harmonized between cohorts. However, C-peptide measurements in different cohorts were calibrated against the same WHO standard, facilitating comparisons. Given the lack of GAD antibody data, it might be suspected that the SIDD group in WellGen includes patients with autoimmune diabetes (Latent Autoimmune Diabetes in Adults: LADA). However, the low prevalence of individuals with high type 1 diabetes GRS scores in the WellGen deems a large contribution of autoimmune diabetes extremely unlikely.

In summary, we demonstrate applicability of a European algorithm for subclassifying type 2 diabetes in young Indian patients. Our results demonstrate a prominent role for insulin secretion defects in the pathophysiology of diabetes in this group. These results have a potential to influence treatment strategies to achieve optimal metabolic control with possible benefits for long-term health [4, 37]. Translation to personalised medicine will come from carefully designed prospective studies including genetic and epigenetic investigations to elucidate pathophysiological mechanisms underlying the subgroups.

## Data Availability

Data is available with Prof C S Yajnik and Dr Rashmi Prasad for sharing to confirm our findings and for additional analyses by applying to the corresponding author with a 200-word plan of analysis. Data sharing is subject to local/regional Institutional Ethics Committees approval and Government of India Health Ministry advisory committee permission.

## Author Contributions

RBP, CSY, and LG contributed to the conception of the work. CSY, SRS and DB contributed to data collection in India, RBP, EA, OA, and LG in Sweden, and AK and TT in Finland. RPB, OA, SRS, EA, RW and PK contributed to the data analysis. RBP, CSY and LG drafted the article. All authors contributed to the interpretation of data and critical revision of the article. All authors gave final approval of the version to be published. RBP and CSY are the guarantors of this work and, as such, had full access to all the data in the study and takes responsibility for the integrity of the data and the accuracy of the data analysis.

## Declaration of interests

We declare no competing interests.

## Acknowledgments

This study was supported by grants from the Indo-Swedish joint network grant from the Swedish Research Council and the Department of Science and Technology, India (Dnr/Reg. nr: 2015-06722) to RBP and C/3019/IFD/2018-2019 to CSY, Wellcome Trust to CSY, Swedish Research Council (2017-02688), Diabetes Wellness Sweden (25-420 PG) and the Novo Nordisk Foundation (NNF18OC0034408) to EA, Swedish Research Foundation (2015-2558) to LG, the Swedish Heart and Lung Foundation, the Crafoord Foundation, the Swedish Diabetes Foundation and the Albert Påhlsson Research Foundation. The DIREVA Study is supported by the Vasa Hospital district, The Academy of Finland (grants no. 263401, 267882, 312063 to LG, 312072 to TT), University of Helsinki, Ollqvist Foundation, Finnish Diabetes Research Foundation, Helsinki University Central Hospital Research Foundation, Jakobstadsnejdens heart foundation to AK. CSY was a visiting Professor to Danish Diabetes Academy (supported by Novo Nordisk Foundation) and Southern University of Denmark during 2016-1019.

We thank all patients and health-care providers for their support and willingness to participate.

We thank Prof Andrew Hattersley, Dr Giriraj Chandak, Dr Shailaja Kale, Dr Sardeshmukh, Dr Meenakumari, Mrs Pallavi Yajnik, Ms Rasika Ladkat, Ms Deepa Raut, Ms Madhura Deshmukh, Ms Chitra Hole for their invaluable contribution to the WellGen Study. We gratefully acknowledge Jacqueline Postma, Johan Hultman, Jasmina Kravic, Gabriella Gremsperger, Ylva Wessman and Ulrika Blom-Nilsson for excellent technical and administrative support; and the ANDIS steering committee for their support.

## Notes

### Competing Interest Statement

The authors have declared no competing interest.

### Author Declarations

All studies were approved by the local/regional Institutional Ethics Committees. The WellGen study was approved by the Institutional Ethics Committee, King Edward Memorial Hospital Research Centre (KEM/HRC/Dir.Off/977, 2004; KEMHRC/LGF/EC/1608A, 2017). The Ahmedabad study was approved by the regional Ethics Committee in Ahmedabad (Rudraksha/Dir.off/380015, 2020). The PHENOEINDY2 study was approved by Institutional Ethics Committee, Assam Medical College (AMC/EC/932, 2007). The ANDIS study was approved by regional ethics review committee in Scania (584/2006 and 2012/676). The DIREVA study was approved by the ethics committee in Vaasa (6/2007). All subjects gave written informed consent.

