## Supplemental_Material for "Subgroups of young type 2 diabetes in India reveal insulin deficiency as a major driver"

#### ESM Methods

##### Comparability of laboratory measurements

C peptide was measured by ELISA (DBC) in the WellGen and Assam study, and by ECLIA (Roche) in the ANDIS and DIREVA study and by CLIA (MAGLUMI) in Ahmedabad study; all based on mouse anti-C-peptide antibody and calibrated against WHO International Reference Reagent for C-peptide of human insulin for immunoassay (IRR code 84/510). This highlights comparability of the assays.

##### GWAS QC and Imputation

SNP exclusion criteria included missingness threshold of > 0.05% MAF<1% and Hardy-Weinberg equilibrium p-value<0.05. Imputation was performed on the Michigan Imputation server using 1000G Phase 3 v5 (GRCh37/hg1) as a reference panel along with Eagle v2.4 phasing and SAS (South Asian) as population type.

### ESM Tables

| | WellGen-Male (902) | | | | | | | | | | | |
|---|---|---|---|---|---|---|---|---|---|---|---|---|
| | Duration of diabetes < 5 y (N=342, 37.9%) | | | | Duration of diabetes ≥ 5y (N=560, 62.1%) | | | | | | | |
| Cluster | SIDD | SIRD | MOD | MARD | SIDD | SIRD | MOD | MARD | P (SIDD) | P (SIRD) | P (MOD) | P (MARD) |
| **Number (%)** | 180 (52.6) | 2 (0.6) | 120 (35.1) | 40 (11.7) | 367 (65.5) | 3 (0.5) | 110 (19.6) | 80 (14.3) | | | | |
| Age at Diagnosis (y) | 37.1 (5.9) | 43.8 (0.21) | 36.9 (5.5) | 41.0 (4.1) | 36.9 (5.5) | 41.2 (3.9) | 36.2 (5.3) | 40.5 (4.1) | 0.726 | 0.440 | 0.281 | 0.570 |
| Duration of diabetes (y) | 1.9 (1.5) | 3.5 (0.27) | 1.9 (1.5) | 2.06 (1.3) | 14.8 (7.5) | 11.06 (7.2) | 14.0 (7.4) | 13.9 (7.9) | -- | -- | -- | -- |
| BMI (Kg/$m^2$) | 24.2 (3.2) | 27.9 (1.6) | 28.1 (3.1) | 24.1 (2.3) | 24.5 (3.0) | 29.3 (4.8) | 29.5 (4.1) | 23.0 (2.3) | 0.208 | 0.715 | **0.003** | 0.015 |
| Fasting glucose ( mmol/L) | 10.1 (2.9) | 5.9 (0.71) | 7.3 (2.1) | 6.7 (1.6) | 10.4 (3.4) | 5.6 (1.3) | 7.9 (2.7) | 6.4 (1.6) | 0.331 | 0.792 | 0.073 | 0.252 |
| HbA1c (mmol/mol) | 87.9 (23.5) | 53.5 (13.1) | 56.7 (11.8) | 46.9 (9.2) | 83.0 (18.8) | 46.1 (9.5) | 63.4 (14.2) | 47.9 (10.4) | 0.008 | 0.507 | **0.0001** | 0.609 |
| Fasting C-Peptide (nmol/L) | 0.73 (0.41) | 2.28 (0.04) | 1.05 (0.45) | 0.70 (0.31) | 0.63 (0.37) | 2.10 (0.62) | 1.12 (0.57) | 0.56 (0.29) | 0.002 | 0.723 | 0.290 | 0.014 |
| HOMA2B | 43.7 (25.1) | 224.0 (43.7) | 94.4 (42.2) | 80.1 (34.9) | 37.2 (20.8) | 232.6 (48.6) | 89.9 (42.9) | 73.3 (30.2) | 0.001 | 0.854 | 0.433 | 0.278 |
| HOMA2IR | 2.06 (1.20) | 5.26 (0.29) | 2.62 (1.27) | 1.71 (0.81) | 1.91 (1.41) | 4.81 (1.69) | 2.90 (1.55) | 1.71 (0.81) | 0.210 | 0.751 | 0.133 | 0.015 |
| | WellGen-Female (710) | | | | | | | | | | | |
| | Duration of diabetes < 5 y (N=238, 33.5%) | | | | Duration of diabetes ≥ 5y (N=472, 66.5%) | | | | | | | |
| Cluster | SIDD | SIRD | MOD | MARD | SIDD | SIRD | MOD | MARD | P (SIDD) | P (SIRD) | P (MOD) | P (MARD) |
| **Number (%)** | 84 (35.3) | 8 (3.4) | 139 (58.4) | 7 (2.9) | 220 (46.6) | 5 (1.1) | 239 (50.6) | 8 (1.7) | | | | |
| Age at Diagnosis (y) | 36.9 (6.3) | 35.4 (4.6) | 37.0 (5.9) | 42.9 (1.1) | 36.6 (5.9) | 40.9 (3.6) | 35.5 (5.9) | 41.2 (3.2) | 0.709 | 0.045 | 0.013 | 0.194 |
| Duration of diabetes (y) | 1.9 (1.5) | 0.99 (1.5) | 2.0 (1.5) | 2.4 (1.9) | 13.6 (6.0) | 13.4 (6.8) | 13.6 (7.1) | 13.9 (6.3) | -- | -- | -- | -- |
| BMI (Kg/$m^2$) | 25.8 (3.8) | 29.6 (7.1) | 28.6 (4.8) | 22.7 (1.4) | 26.2 (3.8) | 25.9 (2.7) | 27.9 (3.8) | 20.7 (3.4) | 0.428 | 0.289 | 0.152 | 0.172 |
| Fasting glucose ( mmol/L) | 11.1 (3.2) | 5.7 (0.85) | 7.5 (1.8) | 6.6 (1.1) | 11.8 (3.6) | 6.2 (2.2) | 8.2 (2.5) | 6.1 (0.9) | 0.122 | 0.551 | **0.003** | 0.292 |
| HbA1c (mmol/mol) | 88.5 (14.5) | 53.4 (12.6) | 57.5 (13.9) | 41.4 (16.7) | 92.3 (16.6) | 69.6 (6.6) | 57.2 (11.7) | 47.8 (8.7) | 0.067 | 0.023 | 0.859 | 0.363 |
| Fasting C-Peptide (nmol/L) | 0.79 (0.38) | 2.0 (0.52) | 0.88 (0.46) | 0.49 (0.19) | 0.69 (0.42) | 1.95 (0.84) | 0.72 (0.39) | 0.35 (0.21) | 0.051 | 0.888 | **0.0001** | 0.188 |
| HOMA2B | 39.2 (22.9) | 217.6 (33.1) | 78.5 (39.9) | 60.2 (17.9) | 32.9 (20.7) | 196.6 (53.5) | 62.7 (37.5) | 58.0 (21.5) | 0.023 | 0.395 | **0.001** | 0.837 |
| HOMA2IR | 2.4 (1.3) | 4.6 (1.3) | 2.2 (1.2) | 1.2 (0.49) | 2.24 (1.54) | 4.7 (2.3) | 1.8 (1.0) | 0.88 (0.43) | 0.343 | 0.947 | **0.004** | 0.192 |

| | WellGen-Total (1612) | | | | | | | | | | | |
|---|---|---|---|---|---|---|---|---|---|---|---|---|
| | Duration of diabetes < 5 y (n=580, 36.0%) | | | | Duration of diabetes ≥ 5y (n=1032, 64.0%) | | | | | | | |
| **Cluster** | **SIDD** | **SIRD** | **MOD** | **MARD** | **SIDD** | **SIRD** | **MOD** | **MARD** | **P (SIDD)** | **P (SIRD)** | **P (MOD)** | **P (MARD)** |
| **Number (%)** | 264 (45.5) | 10 (1.7) | 259 (44.7) | 47 (8.1) | 587 (56.9) | 8 (0.8) | 349 (33.8) | 88 (8.5) | | | | |
| Age at Diagnosis (y) | 37.1 (6.0) | 37.0 (5.4) | 37.0 (5.) | 41.3 (3.9) | 36.8 (5.7) | 41.0 (3.4) | 35.7 (5.7) | 40.6 (4.0) | 0.588 | 0.095 | **0.006** | 0.346 |
| Duration of diabetes (y) | 1.9 (1.5) | 1.5 (1.7) | 1.9 (1.5) | 2.1 (1.4) | 14.3 (7.0) | 12.6 (6.6) | 13.7 (7.2) | 13.9 (7.7) | -- | -- | -- | -- |
| BMI (Kg/m$^2$) | 24.7 (3.5) | 29.3 (6.3) | 28.4 (4.1) | 23.9 (2.3) | 25.1 (3.4) | 28.4 (5.3) | 28.4 (3.9) | 22.8 (2.4) | 0.074 | 0.419 | 0.804 | 0.013 |
| Fasting glucose ( mmol/L) | 10.4 (3.0) | 5.7 (0.79) | 7.4 (1.9) | 6.7 (1.5) | 10.9 (3.5) | 6.0 (1.8) | 8.2 (2.6) | 6.4 (1.4) | 0.047 | 0.700 | **0.0001** | 0.188 |
| HbA1c (mmol/mol) | 88.1 (21.0) | 53.4 (11.9) | 57.1 (12.9) | 46.1 (10.6) | 86.5 (18.5) | 60.9 (14.1) | 59.2 (12.8) | 47.9 (10.3) | 0.257 | 0.247 | 0.054 | 0.338 |
| Fasting C-Peptide (nmol/L) | 0.75 (0.40) | 2.1 (0.47) | 0.96 (0.46) | 0.67 (0.29) | 0.65 (0.39) | 2.0 (0.72) | 0.84 (0.49) | 0.54 (0.28) | 0.001 | 0.856 | **0.004** | 0.014 |
| HOMA2B | 42.2 (24.4) | 218.9 (32.7) | 85.8 (41.7) | 77.1 (33.6) | 35.6 (20.9) | 210.1 (51.57) | 71.3 (41.2) | 71.9 (29.8) | 0.0001 | 0.665 | **0.0001** | 0.361 |
| HOMA2IR | 2.2 (1.3) | 4.7 (1.2) | 2.4 (1.2) | 1.6 (0.79) | 2.03 (1.47) | 4.7 (2.0) | 2.19 (1.31) | 1.31 (0.73) | 0.169 | 0.992 | 0.048 | 0.016 |

*Note: Values are mean (SD), p-value by ANOVA. Bonferroni corrected significant p-values are indicated in bold.*

**ESM table 1: Characteristics of participants enrolled in WellGen study by duration of diabetes and clusters, presented separately for males and females.**

| | Male (902) | | | | Female (710) | | | | Total (1612) | | | |
|---|---|---|---|---|---|---|---|---|---|---|---|---|
| **k-means** | **1** | **2** | **p** | **p1** | **1** | **2** | **p** | **p1** | **1** | **2** | **p** | **p1** |
| **Number (%)** | **590 (65.4)** | **312 (34.6)** | | | **483 (68.0)** | **227 (32.0)** | | | **1073 (66.6)** | **539 (33.4)** | | |
| Age at Diagnosis (y) | 36.66 (5.76) | 38.76 (4.92) | **<0.0001** | **<0.0001** | 35.94 (6.02) | 37.59 (5.77) | **<0.0001** | **<0.0001** | 36.033 (5.89) | 38.27 (5.14) | **<0.0001** | **<0.0001** |
| Duration of diabetes (y) | 10.33 (8.64) | 8.48 (8.21) | **0.0001** | -- | 10.40 (7.53) | 8.00 (7.69) | **0.0001** | -- | 10.36 (8.16) | 8.28 (7.16) | **0.0001** | -- |
| BMI (Kg/$m^2$) | 24.04 (2.94) | 28.05 (3.69) | **<0.0001** | **<0.0001** | 25.99 (3.60) | 29.57 (4.54) | **<0.0001** | **<0.0001** | 24.92 (3.40) | 28.69 (4.14) | **<0.0001** | **<0.0001** |
| Fasting glucose (mmol/l) | 9.51 (3.18) | 8.28 (3.19) | **<0.0001** | **<0.0001** | 10.37 (3.43) | 7.57 (2.64) | **<0.0001** | **<0.0001** | 9.90 (3.32) | 7.98 (2.99) | **<0.0001** | **<0.0001** |
| HbA1c (mmol/mol) | 77.41 (23.27) | 65.30 (20.27) | **<0.0001** | **<0.0001** | 78.09 (21.31) | 57.86 (17.20) | **<0.0001** | **<0.0001** | 77.72 (22.41) | 62.17 (19.45) | **<0.0001** | **<0.0001** |
| Fasting C-Peptide (nmol/l) | 0.55 (0.26) | 1.19 (0.48) | **<0.0001** | **<0.0001** | 0.59 (0.30) | 1.13 (0.51) | **<0.0001** | **<0.0001** | 0.57 (0.28) | 1.16 (0.49) | **<0.0001** | **<0.0001** |
| HOMA2B | 40.87 (22.91) | 92.42 (44.29) | **<0.0001** | **<0.0001** | 36.94 (19.99) | 97.77 (46.21) | **<0.0001** | **<0.0001** | 39.10 (21.72) | 94.67 (45.14) | **<0.0001** | **<0.0001** |
| HOMA2IR | 1.54 (0.80) | 3.19 (1.60) | **<0.0001** | **<0.0001** | 1.78 (1.02) | 2.91 (1.61) | 0.50 | 0.73 | 1.65 (0.91) | 3.07 (1.61) | **<0.0001** | **<0.0001** |

*Note: Values are mean (SD), p-value by ANOVA, p1 adjusted for duration of diabetes. Bonferroni corrected significant p-values are indicated in bold.*

**ESM table 2: Characteristics of participants enrolled in WellGen study by clusters generated using k-means clustering, presented separately for males and females.**

| | WellGen (N=1612) | | | |
|---|---|---|---|---|
| | **1** | **2** | **p-value** | **p1 value** |
| **Number (%)** | **1073 (66.6)** | **539 (33.4)** | | |
| **Current Treatment** | | | | |
| Only on Diet | 68 (6.3) | 67 (12.4) | **0.0001** | **0.0001** |
| Only on OHA (SUs+Metofrmin+/-Glitazones) | 605 (56.4) | 374 (69.4) | | |
| Only on Insulin | 72 (6.7) | 13 (2.4) | | |
| Both OHA+ Insulin[1] | 328 (30.6) | 85 (15.8) | | |
| **Complications** | | | | |
| **Cardiovascular disease[2]** | **76 (7.1)** | **44 (8.2)** | 0.436 | 0.050 |
| Coronary Events | 63 (5.9) | 37 (6.9) | | |
| Stroke | 17 (1.6) | 9 (1.7) | | |
| **Nephropathy (Proteinuria and / or CKD) (n=1612)** | **406 (37.8)** | **155 (28.8)** | **0.0001** | 0.076 |
| **Proteinuria [3]** | 165 (16.0) | 85 (16.5) | 0.813 | 0.271 |
| **CKD: e-Glomerular Filtration Rate*[4] (n=1471)** | **316 (32.7)** | **97 (19.2)** | **0.0001** | **0.0001** |
| Early CKD (60-90) | 249 (25.8) | 81 (16.0) | | |
| Moderate (30-60) | 62 (6.4) | 12 (2.4) | | |
| Severe (<30) | 5 (0.6) | 4 (0.8) | | |
| **Diabetic Retinopathy (n=657) [5]** | **144 (32.1) (n=448)** | **36 (17.2) (n=209)** | **0.0001** | 0.013 |
| NPDR | 135 (30.1) | 32 (15.3) | | |
| PDR | 9 (2.0) | 4 (1.9) | | |
| **Neuropathy [6]** | **449 (43.0)** | **261 (49.2)** | 0.020 | 0.697 |

*Values are number (%). p-value by Chi-square test. * Based on MDRD formula. P1-value by logistic regression adjusted for duration of diabetes and gender. Bonferroni corrected significant p-values are indicated in bold.*

**ESM table 3: Treatment and complications by cluster in the WellGen by clusters generated using k-means clustering.**

| Sl No | SNP | Gene | Major allele | Minor allele | Risk allele | Odds Ratio | Weight |
|---|---|---|---|---|---|---|---|
| 1 | rs2187668 | HLA-DRB1 | C | T | T | - | - |
| 2 | rs7454108 | HLA-DQ8 | C | T | C | - | - |
| | rs3957146 | | C | T | C | - | - |
| 3 | rs1264813 | HLA_A_24 | C | T | T | 1.54 | 0.43 |
| 4 | rs2395029 | HLA_B_5701 | T | G | T | 2.5 | 0.92 |
| 5 | rs3129889 | HLA_DRB1_15 | G | A | A | 14.88 | 2.7 |
| 6 | rs2476601 | PTPN22 | A | G | A | 1.96 | 0.67 |
| 7 | rs689 | INS | A | T | T | 1.75 | 0.56 |
| 8 | rs12722495 | IL2RA | T | C | T | 1.58 | 0.46 |
| 9 | rs2292239 | ERBB3 | G | T | T | 1.35 | 0.3 |

| SNPs | Haplotypes | Haplotypes | Haplotype allele score | Odds ratio | Weights |
|---|---|---|---|---|---|
| rs2187668, rs7454108* (proxy 1: rs3957146) | DR3/DR4 | CT/CT | 6 | 48.18 | 3.87 |
| | DR4/DR4 | CC/CC | 5 | 21.98 | 3.09 |
| | DR3/DR3 | TT/TT | 4 | 21.12 | 3.05 |
| | DR4/X | CC/CT | 3 | 7.03 | 1.95 |
| | DR3/X | CT/TT | 2 | 4.53 | 1.51 |
| | X/X | CC/TT | 1 | 1 | 0 |

*Barker et.al., Winkler et.al.

**ESM table 4. List of type 1 diabetes associated SNPs used to construct genetic risk scores for T1D presented with the risk alleles.**

| | Included (N=560) | Excluded (N=1052) | P-value |
|---|---|---|---|
| Age at Diagnosis (y) | 37.212 (5.832) | 36.863 (5.749) | 0.25 |
| BMI (Kg/m$^2$) | 26.371 (4.028) | 26.079 (4.09) | 0.167 |
| Fasting glucose (mmol/l) | 9.278 (3.265) | 9.251 (3.38) | 0.879 |
| HbA1c (mmol/mol) | 71.271 (22.532) | 73.185 (22.737) | 0.106 |
| Fasting C-Peptide (nmol/l) | 0.748 (0.51) | 0.779 (0.434) | 0.213 |
| HOMA2B | 54.428 (37.141) | 59.419 (42.854) | 0.015 |
| HOMA2IR | 2.102 (1.558) | 2.144 (1.257) | 0.58 |

Values are mean (SD). P value is calculated using T-test. *Bonferroni corrected significant p-values are indicated in bold.*

**ESM table 5: Patient characteristics for those included in the T1D GRS calculation compared to those excluded.**

| | Beta | SE | Z | P |
|---|---|---|---|---|
| **WellGen** | | | | |
| **T1D vs controls (positive control)** | 15.43 | 1.24 | 12.47 | **<2e-16** |
| **SIDD vs controls** | -1.38 | 1.07 | -1.29 | 0.197 |
| **MOD vs controls** | -0.90 | 1.08 | -0.83 | 0.40 |
| **T1D vs SIDD** | 14.30 | 1.33 | 10.73 | **<2e-16** |
| **T1D vs MOD** | 13.01 | 1.31 | 9.93 | **<2e-16** |

*Bonferroni corrected significant p-values are indicated in bold.*

**ESM table 6. Association of type 1 diabetes genetic risk scores with T1D (as positive control), SIDD, and MOD clusters.**

| | DIREVA | | |
|---|---|---|---|
| **Characteristics** | **Male** | **Female** | **All** |
| Number | 243 | 177 | 420 |
| Age at Diagnosis (y) | 36.78 (7.23) | 36.44 (6.95) | 36.6 (7.1) |
| Duration of Diabetes (y) | 14.78 (12.06) | 13.85 (12.30) | 14.39 (12.16) |
| BMI ($Kg/m^2$) | 31.62 (6.59) | 31.63 (6.60) | 31.62 (6.6) |
| Fasting glucose (mmol/L) | 9.19 (3.06) | 8.74 (3.25) | 9.01 (3.15) |
| HbA1c (mmol/mol) | 59.32 (16.93) | 56.48 (16.84) | 58.13 (16.93) |
| Fasting C-peptide (nmol/L) | 0.60 (0.51) | 0.59 (0.41) | 0.59 (0.47) |
| HOMA2-B | 46.80 (36.26) | 53.61 (36.17) | 49.67 (34.09) |
| HOMA2-IR | 1.73 (1.36) | 1.65 (1.03) | 1.69 (1.23) |

*Note: Values are mean (SD), pvalue calculated using t-test*
* Follow up time.

**ESM table 7: Clinical and Biochemical characteristics of participants enrolled in DIREVA study with age at diagnosis less than 45years and comparison with WellGen participants.**

| | DIREVA | | | | | | WellGen vs DIREVA | |
|---|---|---|---|---|---|---|---|---|
| **Gender** | **All (420)** | | | | | | | |
| **Cluster** | **SIDD** | **SIRD** | **MOD** | **MARD** | **p** | **p1** | **p: SIDD** | **p:MOD** |
| **Number (%)** | 99 (23.57) | 1 (3.0) | 297 (70.7) | 23 (5.47) | | | | |
| Age at Diagnosis (y) | 34.30 (7.70) | 41.20 (0) | 36.99 (6.88) | 41.94 (2.16) | **< 0.0001** | **< 0.0001** | **<0.0001** | 0.08 |
| Duration of diabetes (y) | 20.48 (13.26) | 0.35 (6.46) | 12.39 (11.06) | 14.54 (12.54) | | | | |
| BMI ($Kg/m^2$) | 27.03 (3.97) | 37.80 (0) | 33.65 (6.42) | 25.00 (1.97) | **< 0.0001** | **< 0.0001** | **< 0.0001** | **< 0.0001** |
| Fasting glucose (mmol/L) | 11.10 (3.7) | 5.9 (0) | 8.42 (2.65) | 7.66 (2.09) | **< 0.0001** | **< 0.0001** | 0.69 | **0.001** |
| HbA1c (mmol/mol) | 75.31 (16.74) | 38 (0) | 53.47 (13.24) | 45.25 (6.43) | **< 0.0001** | **< 0.0001** | **< 0.0001** | **< 0.0001** |
| Fasting C-Peptide (nmol/L) | 0.32 (0.30) | 2.27 (0) | 0.70 (0.48) | 0.36 (0.2) | **< 0.0001** | **< 0.0001** | | |
| HOMA2B | 24.22 (15.64) | 219.3 (0) | 58.21 (33.85) | 41.64 (18.63) | **< 0.0001** | **< 0.0001** | | |
| HOMA2IR | 1.23 (0.96) | 5.23 (0) | 1.89 (1.27) | 0.97 (0.47) | **< 0.0001** | **< 0.0001** | | |

| **Gender** | **Male (243)** | | | | | | | |
|---|---|---|---|---|---|---|---|---|
| **Cluster** | **SIDD** | **SIRD** | **MOD** | **MARD** | **p** | **p1** | **p: SIDD** | **p:MOD** |
| **Number (%)** | 77 (31.69) | - | 148 (60.91) | 18 (7.4) | | | | |
| Age at Diagnosis (y) | 33.94 (7.86) | - | 37.66 (6.76) | 48.21 (3.76) | **<0.0001** | **<0.0001** | **<0.0001** | 0.09 |
| Duration of diabetes (y) | 20.77 (13.62) | - | 11.74 (10.20) | 13.64 (8.61) | | | | |
| BMI ($Kg/m^2$) | 26.85 (3.53) | - | 34.85 (6.11) | 32.30 (10.42) | **<0.0001** | **<0.0001** | **<0.0001** | **<0.0001** |
| Fasting glucose ( mmol/L) | 10.59 (3.55) | - | 8.63 (2.60) | 9.08 (3.14) | **<0.0001** | **<0.0001** | 0.45 | **0.0002** |
| HbA1c (mmol/mol) | 72.47 (16.55) | - | 54.18 (13.61) | 45.35 (6.09) | **<0.0001** | **<0.0001** | **<0.0001** | **0.0001** |
| Fasting C-Peptide (nmol/L) | 0.32 (0.28) | - | 0.78 (0.54) | 0.65 (0.39) | **<0.0001** | **<0.0001** | | |
| HOMA2B | 25.52 (15.73) | - | 58.59 (34.06) | 49.50 (23.62) | **<0.0001** | **<0.0001** | | |
| HOMA2IR | 1.15 (0.86) | - | 2.12 (1.50) | 1.84 (1.02) | **<0.0001** | **<0.0001** | | |

| Gender | Female (177) | | | | | | | |
|---|---|---|---|---|---|---|---|---|
| Cluster | SIDD | SIRD | MOD | MARD | p | p1 | p: SIDD | p:MOD |
| **Number (%)** | 22 (12.43) | 1 (0.56) | 149 (84.18) | 5 (2.82) | | | | |
| Age at Diagnosis (y) | 35.54 (7.10) | 41.20(0) | 36.33 (6.96) | 42.98 (1.95) | 0.088 | 0.084 | 0.40 | 0.24 |
| Duration of diabetes (y) | 19.49(12.15) | 0.35 (-) | 13.04 (11.84) | 15.89 (21.23) | | | | |
| BMI (Kg/m$^2$) | 27.67 (5.30) | 37.80 (0) | 32.46 (6.50) | 23.28 (0.87) | **<0.0001** | **<0.0001** | 0.06 | 0.24 |
| Fasting glucose ( mmol/L) | 12.90 (3.92) | 5.90 (0) | 8.20 (2.69) | 6.96 (1.90) | **<0.0001** | **<0.0001** | 0.10 | 0.35 |
| HbA1c (mmol/mol) | 85.23 (13.56) | 38 (0) | 52.75 (12.87) | 44.87 (8.35) | **<0.0001** | **<0.0001** | 0.09 | **0.0002** |
| Fasting C-Peptide (nmol/L) | 0.34 (0.32) | 2.27 (0) | 0.63 (0.39) | 0.28 (0.21) | **0.0015** | **0.0016** | | |
| HOMA2B | 19.72 (14.81) | 219.3 (0) | 57.82 (33.76) | 44.30 (17.39) | **<0.0001** | **<0.0001** | | |
| HOMA2IR | 1.52 (1.26) | 5.23 (0) | 1.67 (0.96) | 0.82 (0.40) | **<0.0001** | **<0.0001** | | |

*Note: Values are mean (SD), p-value by ANOVA, p1 adjusted for duration of diabetes. P: SIDD and P: MOD= p values for SIDD group and MOD group comparisons between WellGen and DIREVA respectively. Bonferroni corrected significant p-values are indicated in bold.*

**ESM table 8: Characteristics of participants enrolled in the DIREVA study by clusters for all participants, males and females.**

| | Ahmedabad | | |
|---|---|---|---|
| **Characteristics** | **Male** | **Female** | **All** |
| Number | 138 (73.79) | 49 (26.20) | 187 |
| Age at Diagnosis (y) | 36.6 (6.75) | 37.47 (6.72) | 36.83 (6.74) |
| BMI (Kg/m$^2$) | 27.44 (3.7) | 28.28 (4.16) | 27.66 (3.83) |
| Fasting glucose (mmol/L) | 10.70 (4.64) | 11.37 (4.78) | 10.87 (4.68) |
| HbA1c (mmol) | 72.61 (26.2) | 72.91 (25) | 72.69 (25.82) |
| Fasting C-peptide (nmol/L) | 0.45 (0.21) | 0.41 (0.16) | 0.44 (0.20) |
| HOMA2-B | 37.41 (31.54) | 30.35 (23.94) | 35.56 (29.84) |
| HOMA2-IR | 1.5 (1.1) | 1.48 (1.01) | 1.49 (1.07) |

**ESM table 9: Clinical and Biochemical characteristics of participants enrolled in Ahmedabad study with age at diagnosis less than 45 years**

| | Ahmedabad | | | | |
|---|---|---|---|---|---|
| **Gender** | **All (187)** | | | | |
| **Cluster** | **SIDD** | **SIRD** | **MOD** | **MARD** | **p** |
| **Number (%)** | 106 (56.68) | 0 (0) | 62 (33.15) | 19 (10.16) | |
| Age at Diagnosis (y) | 37.05(6.66) | | 34.84 (6.79) | 42.05(3.34) | **0.0002** |
| BMI (Kg/$m^2$) | 26.76(3.34) | | 29.99 (3.79) | 25.06(2.68) | **<0.0001** |
| Fasting glucose ( mmol/L) | 13.03 (4.53) | | 8.48 (3.26) | 6.66 (2.13) | **<0.0001** |
| HbA1c (mmol) | 88.25(22.94) | | 53.6 (11.18) | 48.29(8.91) | **<0.0001** |
| Fasting C-Peptide (nmol/L) | 0.43 (0.20) | | 0.48 (0.18) | 0.44 (0.20) | 0.282 |
| HOMA2B | 22.83(18.63) | | 48.48 (30.84) | 64.38(39.32) | **<0.0001** |
| HOMA2IR | 1.66(1.22) | | 1.34(0.86) | 1.06(0.52) | 0.032 |
| **Gender** | **Ahmedabad: Male (138)** | | | | |
| **Cluster** | **SIDD** | **SIRD** | **MOD** | **MARD** | **p** |
| **Number** | 85 (61.59) | 0 (0) | 36 (26.08) | 17 (12.32) | |
| Age at Diagnosis (y) | 36.63(6.85) | | 34.11(6.44) | 41.71(3.37) | **<0.0001** |
| BMI (Kg/$m^2$) | 26.5(3.32) | | 30.66 (2.97) | 25.33(2.68) | **<0.0001** |
| Fasting glucose ( mmol/L) | 12.66 (4.45) | | 8.05 (3.24) | 6.50 (1.71) | **<0.0001** |
| HbA1c (mmol) | 86.38(23.16) | | 52.09 (12.55) | 47.22(8.79) | **<0.0001** |
| Fasting C-Peptide (nmol/L) | 0.43 (0.22) | | 0.51 (0.18) | 0.45 (0.21) | 0.218 |
| HOMA2B | 24.17(19.34) | | 56.15 (35.34) | 63.91(36.8) | **<0.0001** |
| HOMA2IR | 1.62(1.23) | | 1.4 (0.91) | 1.1(0.54) | 0.168 |
| **Gender** | **Ahmedabad: Female (49)** | | | | |
| **Cluster** | **SIDD** | **SIRD** | **MOD** | **MARD** | **p** |
| **Number (%)** | 21 (42.86) | 0 (0) | 26 (53.06) | 2 (4.10) | |

| | | | | | |
|---|---|---|---|---|---|
| Age at Diagnosis (y) | 38.76(5.66) | | 35.85 (7.24) | 45(0) | 0.084 |
| BMI (Kg/m$^2$) | 27.83(3.31) | | 29.07 (4.59) | 22.76(1.68) | 0.093 |
| Fasting glucose ( mmol/L) | 14.53 (4.66) | | 8.89 (3.34) | 7.99 (5.57) | **<0.0001** |
| HbA1c (mmol) | 95.79(20.86) | | 55.63 (9.05) | 57.38(3.09) | **<0.0001** |
| Fasting C-Peptide (nmol/L) | 0.40 (0.14) | | 0.44 (0.18) | 0.31 (0.05) | 0.480 |
| HOMA2B | 17.41(14.6) | | 37.87 (19.23) | 68.35(78.28) | **<0.0001** |
| HOMA2IR | 1.82(1.19) | | 1.25 (0.8) | 0.75(0.06) | 0.094 |

*Note: Values are mean (SD), p-value by ANOVA. Bonferroni corrected significant p-values are indicated in bold.*

#### ESM table 10. Characteristics of participants enrolled in the Ahmedabad study by clusters for all participants, for males and females

| | Assam (n=205) | | |
|---|---|---|---|
| **Characteristics** | **Male** | **Female** | **All** |
| Number | 135 (65.85) | 70 (34.15) | 205 |
| Age at Diagnosis (y) | 32.93 (4.86) | 30.99 (5.8) | 32.26 (5.27) |
| Duration of Diabetes (y) | 2.87 (3.38) | 4.33 (3.85) | 3.37 (3.60) |
| BMI (Kg/m$^2$) | 23.46 (3.86) | 23.21 (3.65) | 23.37 (3.79) |
| Fasting glucose (mmol/L) | 10.32 (4.85) | 10.90 (4.93) | 10.52 (4.87) |
| HbA1c (mmol) | 82.3 (33.38) | 82.15 (30.98) | 82.25 (32.5) |
| Fasting C-peptide (nmol/L) | 0.40 (0.31) | 0.45 (0.38) | 0.42 (0.34) |
| HOMA2-B | 38.96 (35.35) | 34.93 (28.45) | 37.58 (33.14) |
| HOMA2-IR | 1.55 (2.71) | 2.26 (4.98) | 1.79 (3.65) |

**ESM table 11: Clinical and Biochemical characteristics of participants enrolled in Assam study with age at diagnosis less than 45years**

| | **Assam** | | | | | |
|---|---|---|---|---|---|---|
| **Gender** | **All (205)** | | | | | |
| **Cluster** | **SIDD** | **SIRD** | **MOD** | **MARD** | **p** | **p1** |
| **Number (%)** | 138 (66.66) | 3 (1.40) | 48 (23.20) | 16 (7.72) | | |
| Age at Diagnosis (y) | 32.14 (5.3) | 34 (6.56) | 31.48 (5.44) | 35.38 (3.1) | 0.069 | 0.112 |
| Duration of diabetes (y) | 3.66 (3.83) | 4.26 (5.04) | 3.01 (3.15) | 1.81 (2.00) | 0.209 | - |
| BMI (Kg/$m^2$) | 22.68 (3.43) | 17.6 (3.5) | 26.25 (3.44) | 21.79 (2.95) | **<0.0001** | **<0.0001** |
| Fasting glucose ( mmol/L) | 11.89 (4.14) | 29.11 (3.32) | 6.84 (2.08) | 6.18 (1.27) | **<0.0001** | **<0.0001** |
| HbA1c (mmol) | 97.06 (26.51) | 105.46 (52.95) | 52.44 (13.91) | 39.62 (7.9) | **<0.0001** | **<0.0001** |
| Fasting C-Peptide (nmol/L) | 0.38 (0.32) | 0.07 (0.01) | 0.57 (0.39) | 0.33 (0.25) | **0.002** | **0.002** |
| HOMA2B | 24.26 (18.7) | 3.2 (0) | 71.03 (39.28) | 58.6 (31.49) | **<0.0001** | **<0.0001** |
| HOMA2IR | 1.4 (1.19) | 30.3 (0) | 1.45 (0.92) | 0.88 (0.52) | **<0.0001** | **<0.0001** |
| **Gender** | **Assam: Male (135)** | | | | | |
| **Cluster** | **SIDD** | **SIRD** | **MOD** | **MARD** | **p** | **p1** |
| **Number (%)** | 99 (73.33) | 1 (0.74) | 21 (15.55) | 14 (10.37) | | |
| Age at Diagnosis (y) | 32.53 (4.93) | 40 (NA) | 32.86 (4.96) | 35.36 (3.32) | 0.096 | 0.225 |
| Duration of diabetes (y) | 3.20 (3.57) | 0.15 (NA) | 2.23 (3.06) | 1.72 (1.96) | 0.267 | - |
| BMI (Kg/$m^2$) | 22.73(3.53) | 15.35 (NA) | 28.03(2.44) | 22.39(2.52) | **<0.0001** | **<0.0001** |
| Fasting glucose ( mmol/L) | 11.59 (4.35) | 31.97 (NA) | 5.98 (1.30) | 6.29 (1.29) | **<0.0001** | **<0.0001** |
| HbA1c (mmol) | 94.72(27.32) | 163.39 (NA) | 48.22(13.56) | 39.81(8.46) | **<0.0001** | **<0.0001** |
| Fasting C-Peptide (nmol/L) | 0.36 (0.28) | 0.08 (NA) | 0.64 (0.40) | 0.35 (0.26) | **0.001** | **0.001** |
| HOMA2B | 25.32(19.57) | 3.2 (NA) | 91.66(39.73) | 58.89(33.75) | **<0.0001** | **<0.0001** |
| HOMA2IR | 1.35(1.14) | 30.3 (NA) | 1.56(0.9) | 0.92(0.54) | **<0.0001** | **<0.0001** |

| Gender | Assam: Female (70) | | | | | |
|---|---|---|---|---|---|---|
| **Cluster** | **SIDD** | **SIRD** | **MOD** | **MARD** | **p** | **p1** |
| **Number (%)** | 39 (54.16) | 2 (2.77) | 27 (37.5) | 2 (2.77) | | |
| Age at Diagnosis (y) | 31.15 (6.1) | 31 (5.66) | 30.41 (5.65) | 35.5 (0.71) | 0.689 | 0.562 |
| Duration of diabetes (y) | 4.82 (4.27) | 6.32 (5.04) | 3.61 (3.14) | 2.44 (3.02) | 0.467 | - |
| BMI (Kg/m$^2$) | 22.58 (3.19) | 18.73 (4.11) | 24.86 (3.49) | 17.61 (2.79) | **0.001** | **0.002** |
| Fasting glucose ( mmol/L) | 12.68 (3.50) | 27.69 (3.13) | 7.50 (2.34) | 5.41 (1.06) | **<0.0001** | **<0.0001** |
| HbA1c (mmol) | 103 (23.61) | 76.5 (23.96) | 55.72 (13.52) | 38.25 (0.77) | **<0.0001** | **<0.0001** |
| Fasting C-Peptide (nmol/L) | 0.44 (0.40) | 0.07 (0) | 0.51 (0.38) | 0.21 (0.18) | 0.333 | 0.381 |
| HOMA2B | 21.56 (16.2) | 3.2 (0) | 54.99 (31.04) | 56.55 (7.28) | **<0.0001** | **<0.0001** |
| HOMA2IR | 1.53 (1.32) | 30.3 (0) | 1.37 (0.95) | 0.62 (0.26) | **<0.0001** | **<0.0001** |

*Note: Values are mean (SD), p-value by ANOVA, p1 adjusted for duration of diabetes. Bonferroni corrected significant p-values are indicated in bold.*

**ESM Table 12. Characteristics of participants enrolled in the Assam study by clusters for all participants, males and females.**

#### ESM Figures

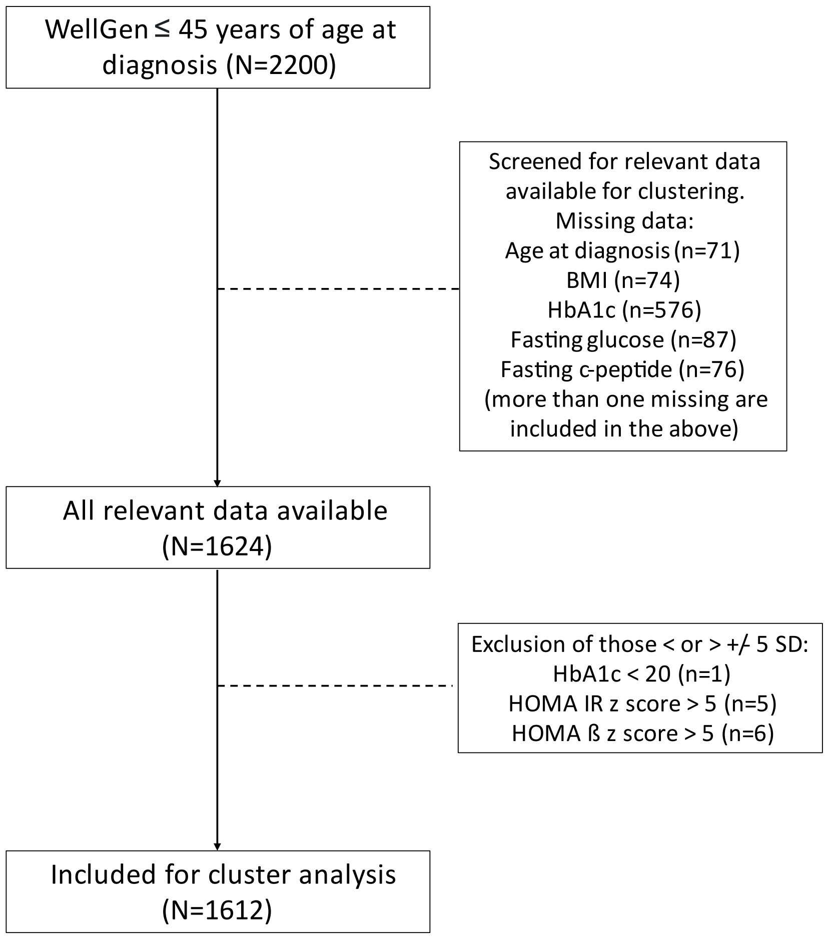

**ESM figure 1: Flowchart for the WellGen cohort.**

**2a**

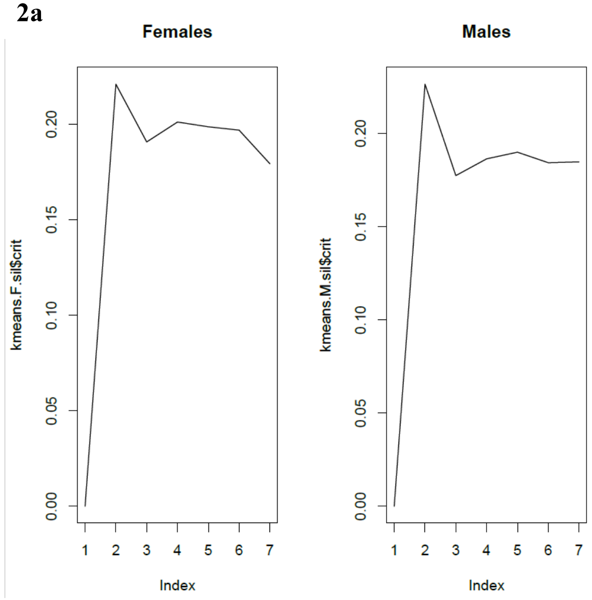

**2b**

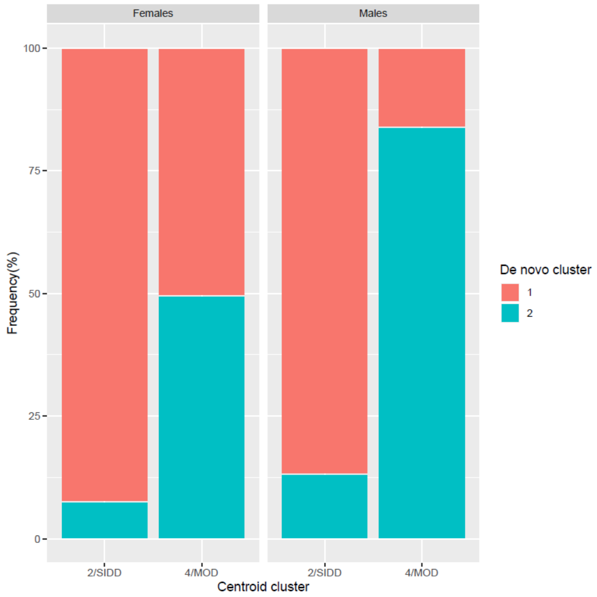

**ESM figure 2.** 2a. Silhouette plot showing optimal number of clusters in the WellGen cohort. 2b. Nearest centroid vs k-means clustering: Overlap of distribution of patients in the two largest clusters derived using two methods. De novo k-means clustering showed two clusters, with patients in cluster 1 showing as ~90 % overlap with those in SIDD (ALL: 89.88%, Males: 86.44%, Females: 96.68%) whereas >patients in cluster 2 patients showed a >70% overlap with MOD (ALL: 72.44%, Males: 89.73% and Females: 61.81%).

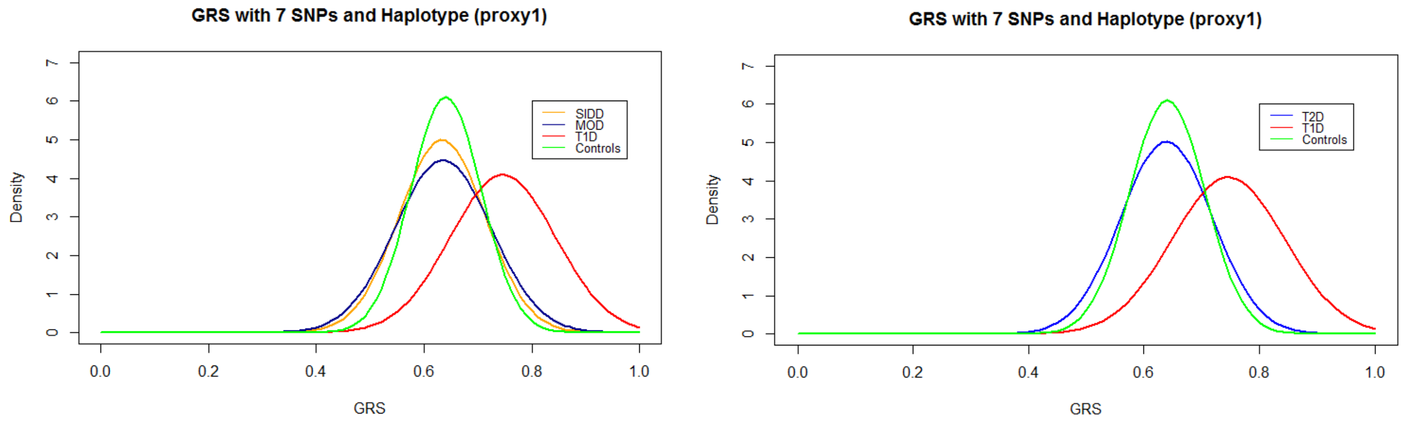

**ESM figure 3**. Density plot of genetic risk score distributions in T1D, SIDD, MOD and controls in the WellGen study.

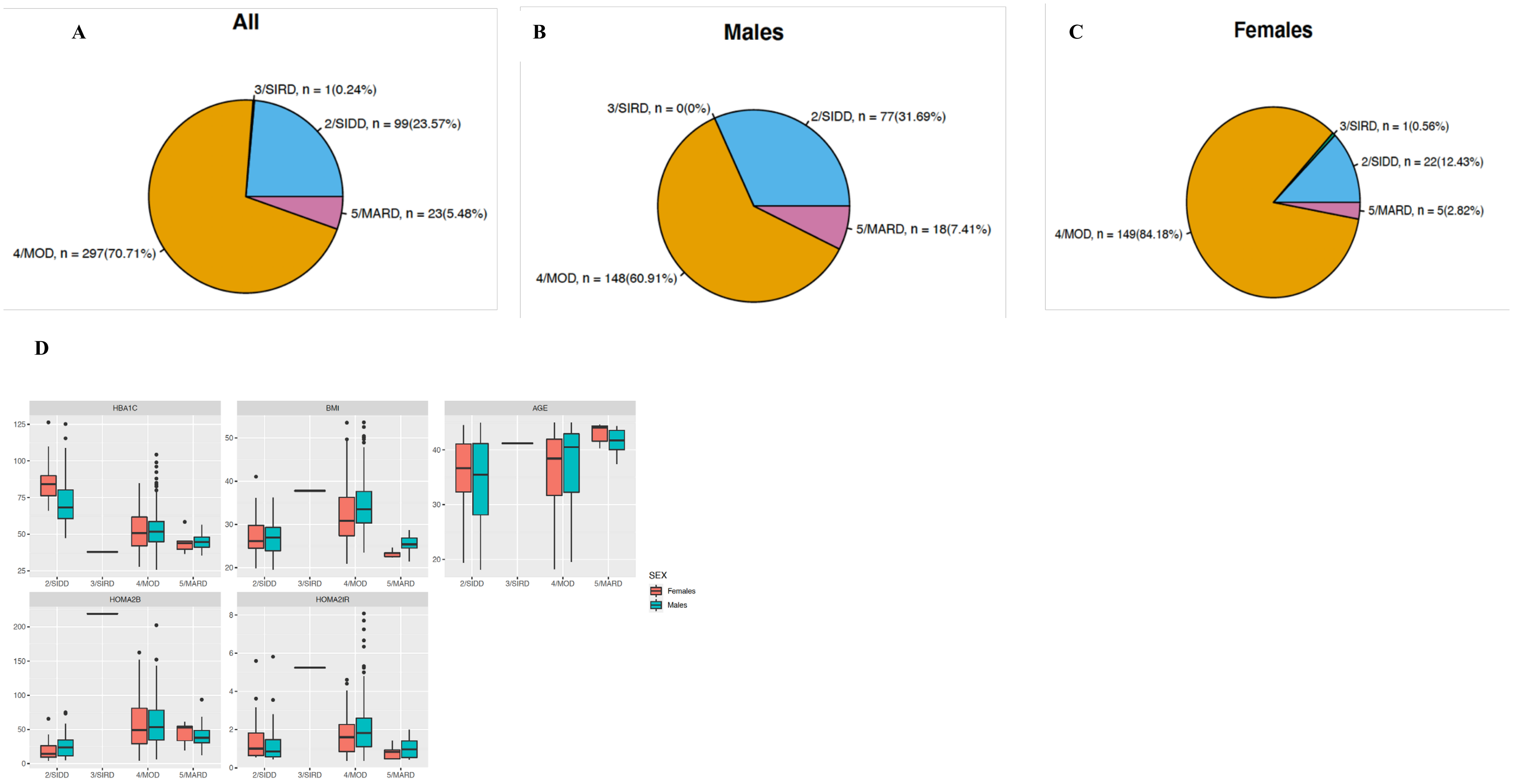

**ESM figure 4**. Patient distribution in various predefined clusters. (A) Distribution of DIREVA patients (n = 424) (B) distribution of men with diabetes from the DIREVA study (n = 243) (C) distribution of women with diabetes from the DIREVA study (n =177). (D) Cluster Characteristics in the DIREVA study. Distribution of age at diagnosis, BMI, HbA1c, HOMA2-B and HOMA2-IR in the DIREVA study for each cluster. k-means clustering was done separately for men and women; data is shown for each sex separately. SIDD = severe insulin-deficient diabetes, SIRD = resistant diabetes, MOD = mild obesity-related-related diabetes, MARD = mild age-related diabetes. HOMA2-B = homeostatic model assessment 2 estimates of beta cell function. HOMA2-B = homeostatic model assessment 2 estimates insulin resistance.

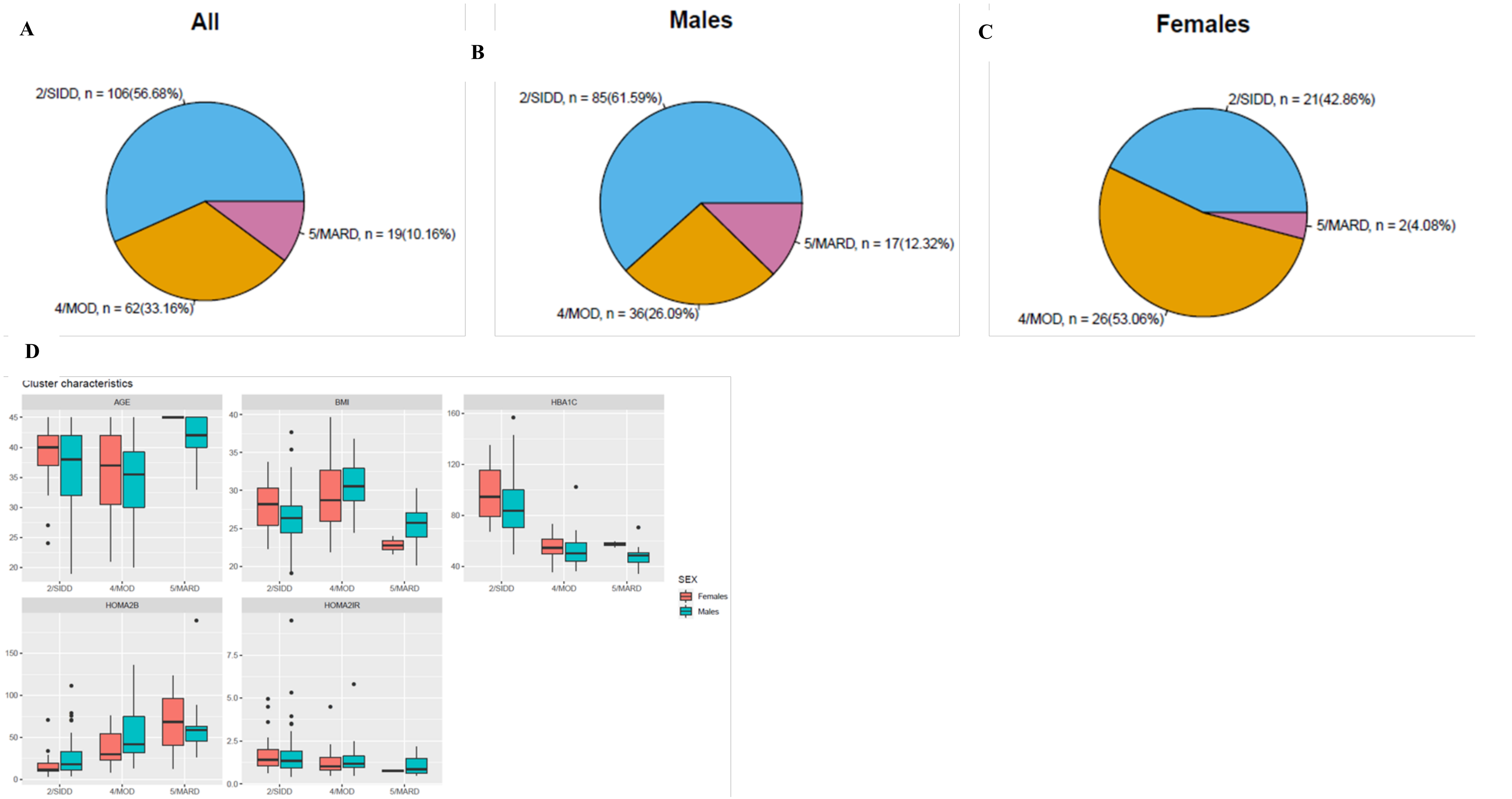

**ESM figure 5**. Distribution of participants from the Ahmedabad study in the predefined clusters. (A) Distribution of patients (n =) (B) distribution of women with diabetes (n =) (C) distribution of men with diabetes (n =) (D) Cluster Characteristics in the Ahmedabad cohort. Distribution of age at diagnosis, BMI, HbA1c, HOMA2-B and HOMA2-IR in the Ahmedabad study for each cluster. k-means clustering was done separately for men and women; data is shown for each sex separately. SIDD = severe insulin-deficient diabetes, SIRD = resistant diabetes, MOD = mild obesity-related-related diabetes, MARD = mild age-related diabetes. HOMA2-B = homeostatic model assessment 2 estimates of beta cell function. HOMA2-B = homeostatic model assessment 2 estimates insulin resistance.

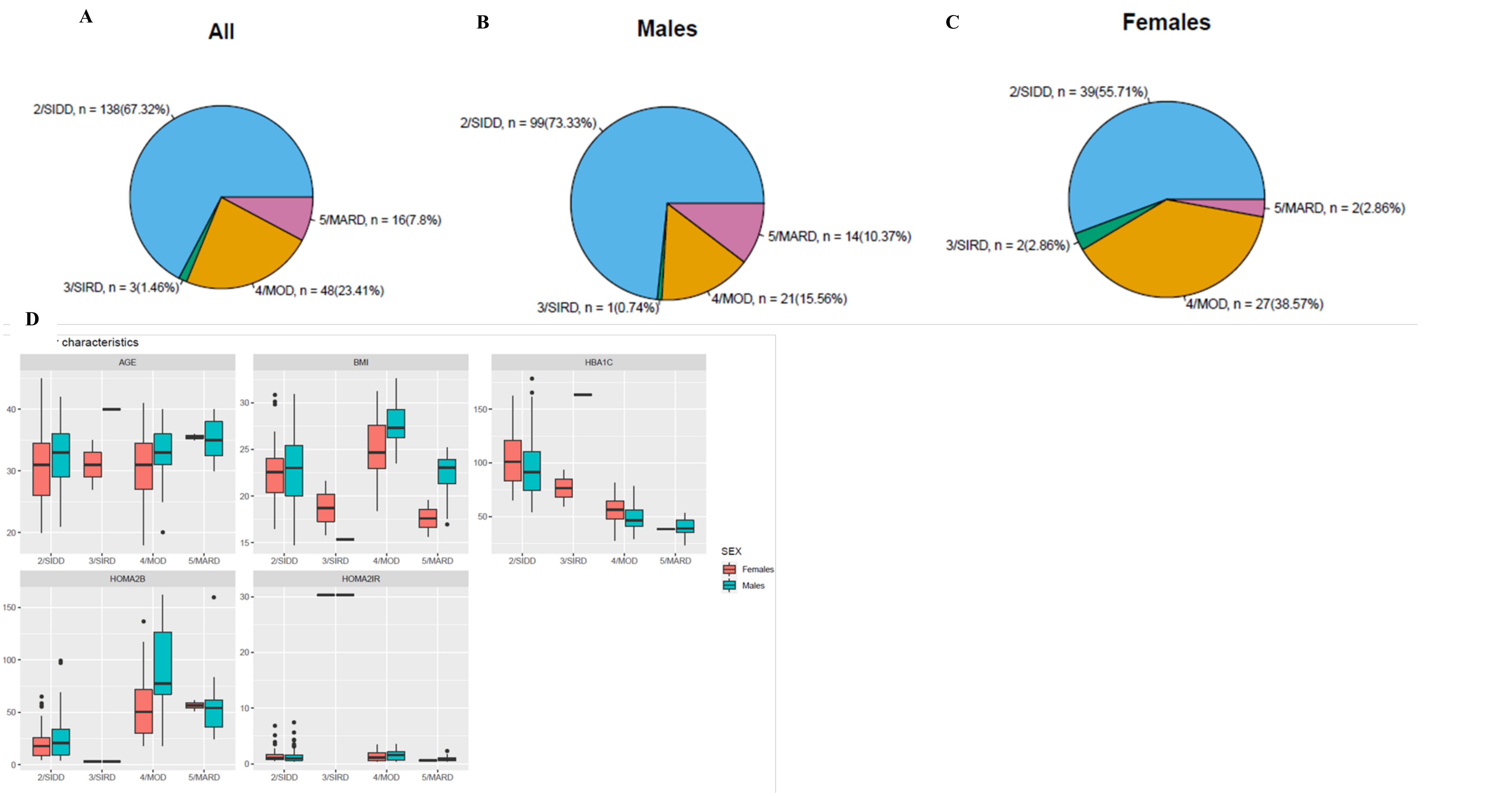

**ESM Figure 6**. Distribution of participants from the Assam study in the predefined clusters. (A) Distribution of patients (n =) (B) distribution of women with diabetes (n =) (C) distribution of men with diabetes (n =) (D) Cluster Characteristics in the Assam cohort. Distribution of age at diagnosis, BMI, HbA1c, HOMA2-B and HOMA2-IR in the Assam study for each cluster. k-means clustering was done separately for men and women; data is shown for each sex separately. SIDD = severe insulin-deficient diabetes, SIRD = resistant diabetes, MOD = mild obesity-related-related diabetes, MARD = mild age-related diabetes. HOMA2-B = homeostatic model assessment 2 estimates of beta cell function. HOMA2-B = homeostastic model assessment 2 estimates insulin resistance.
